# Bipolar Disorder and Education: Deciphering a Complex Relationship using Genetic and Sibling Differences in a Multi-cohort Study

**DOI:** 10.64898/2026.11.13.26368023

**Authors:** Vaishnavi K Madhavan, Perline Demange, Ziada Ayorech, Jouke Jan Hottenga, Meike Bartels, Eivind Ystrom, Michel G Nivard, Wonuola A Akingbuwa

## Abstract

**Background:** While bipolar disorder has a positive genetic correlation (r∼0.3) with educational attainment (EA), individuals with bipolar disorder diagnoses have lower EA in adulthood. To clarify this paradox, we hypothesise that bipolar disorder and educational outcomes share a positive genetic association, reflected in high academic performance earlier in life, but that emerging bipolar disorder symptoms disrupt academic trajectories, leading to the observed poor long-term educational outcomes.

**Methods:** We employed a sibling comparison design across four population-based cohorts (N_total_ = 1,145,795 siblings). First, we tested the association between bipolar disorder polygenic scores (PGS) and educational outcomes (EA and academic performance) in the Norwegian Mother, Father and Child Cohort Study (MoBa) and UK Biobank (UKB). Second, we examined associations between academic performance at ages 10, 12, 13, and 14 and subsequent bipolar disorder diagnosis using registry data from Statistics Netherlands and Statistics Norway (with the Norwegian Control and Payment of Health Reimbursements Databases).

**Findings:** Siblings with higher bipolar disorder PGS had significantly higher EA (β_W_ = 0.03, 95% CI [0.01, 0.05], N=16,746) as well as higher academic performance at age 10 (0.03 [0.01, 0.05], N=17,677) in MoBa. Secondly, individuals with bipolar disorder had better overall academic performance than their unaffected sibling at ages 10 (HR_W_ = 1.12 [1.02, 1.22], N=343,898) and 12 (1.59 [1.16, 2.19], N=619,796). However, individuals with bipolar disorder experienced a decline in academic performance at ages 13 (0.84 [0.73, 0.96], N=320,342) and 14 (0.72 [0.61, 0.85], N=299,039).

**Interpretation:** Bipolar disorder shares a positive genetic association with educational outcomes, reflected in early academic advantage. This is followed by an academic decline in mid-adolescence, highlighting the period of educational disruption associated with bipolar disorder.

**Funding:** The Dutch Ministry of Education, Culture and Sciences, European Research Council, The Research Council of Norway, The Swedish Research Council

## Introduction

Bipolar disorder is associated with significant psychosocial, occupational and socioeconomic disability.^1–3^ This burden extends to educational settings, as evidenced by high levels of school dropout,^4^ lower likelihood of completing a university degree,^5^ and ultimately, lower educational attainment (EA) compared to the general population.^6^

Paradoxically, the rise of behavioural genetics research has consistently demonstrated a positive genetic association between bipolar disorder and educational outcomes, implying a shared genetic liability.^6–11^ For example, EA is shown to be positively associated with bipolar disorder polygenic scores (PGS).^12–15^ A PGS is a single value estimate of an individual’s probabilistic genetic risk for a phenotype, calculated as a sum of genetic variants, weighted by their corresponding effect sizes derived from summary statistic data.^16^ These findings imply that on average, individuals with a higher probabilistic genetic risk for bipolar disorder are predicted to have better educational outcomes.

Consistent with the positive genetic association, studies report high premorbid cognitive abilities in childhood and adolescence.^17–21^ Similarly, there is evidence for high academic performance during adolescence; one registry-based Swedish study reports that individuals with high school grades at age 15 had a fourfold risk of bipolar disorder compared to those with average grades.^22^ However, evidence for elevated academic performance is not very consistent, with studies from Finland and Denmark reporting null or even negative associations between bipolar disorder and school performance at ages 15 - 17.^23–25^

To date, contradictions between the genetic and phenotypic associations have not been addressed. On one hand, the positive associations between bipolar disorder and educational outcomes could reflect population-level confounding like socioeconomic strata (SES). For instance, individuals from highly-educated and high SES families are more likely to be diagnosed with bipolar disorder,^6,26^ potentially resulting in a positive association with educational outcomes due to gene-environment correlation.

Alternatively, a true positive genetic correlation between bipolar disorder and educational outcomes may exist, yet only partially expressed at the phenotypic level, with associations varying across developmental stages. However, all previous studies have examined cross-sectional associations with academic performance measured at a single age, limiting insight into the longitudinal trajectory and timing of educational disruption.^22–25^ Monitoring the academic progress of those with bipolar disorder across development may point to clinically and scientifically salient early processes related to the prodromal phase of bipolar disorder. A better understanding of the early prodromal phase is critical given that bipolar disorder typically involves a significant period of diagnostic delay, with depressive symptoms first emerging in early adolescence and a diagnosis of bipolar disorder provided at ages 25– 30 on average.^27^

To clarify the paradoxical associations between bipolar disorder and educational outcomes, we tested a model which posits that the genetic liability for bipolar disorder is positively associated with the genetic liability for educational outcomes – both early-life academic performance and long-term EA, represented by the green pathways in **Figure 1a**. This shared genetic association is reflected in an (initial) academic benefit, as indicated by the positive association with early-life academic performance (pink pathway). However, if an individual develops bipolar disorder, their academic progress may be hindered, ultimately leading to a negative association between the disorder and long-term educational outcomes such as EA, as evidenced previously (the blue pathway).^6,28^

We performed two analyses across four different cohorts in the UK, Norway, and the Netherlands, using a genetically-informed sibling comparison design to account for unmeasured familial confounding shared between siblings raised in a single household.^29^ First, to confirm a positive genetic association between bipolar disorder and educational outcomes, we examined the within-sibship association between bipolar disorder PGS and each of our two educational outcomes: EA and academic performance (analysis 1). Second, we tested the within-sibship association between academic performance across ages 10, 12, 13, and 14 and subsequent bipolar disorder diagnosis (analysis 2). This longitudinal investigation of academic performance from ages 10 to 14 enabled us to monitor the academic trajectory of individuals with bipolar disorder typically before the average age of diagnosis and potentially identify the period of educational disruption.

**Figure 1.**
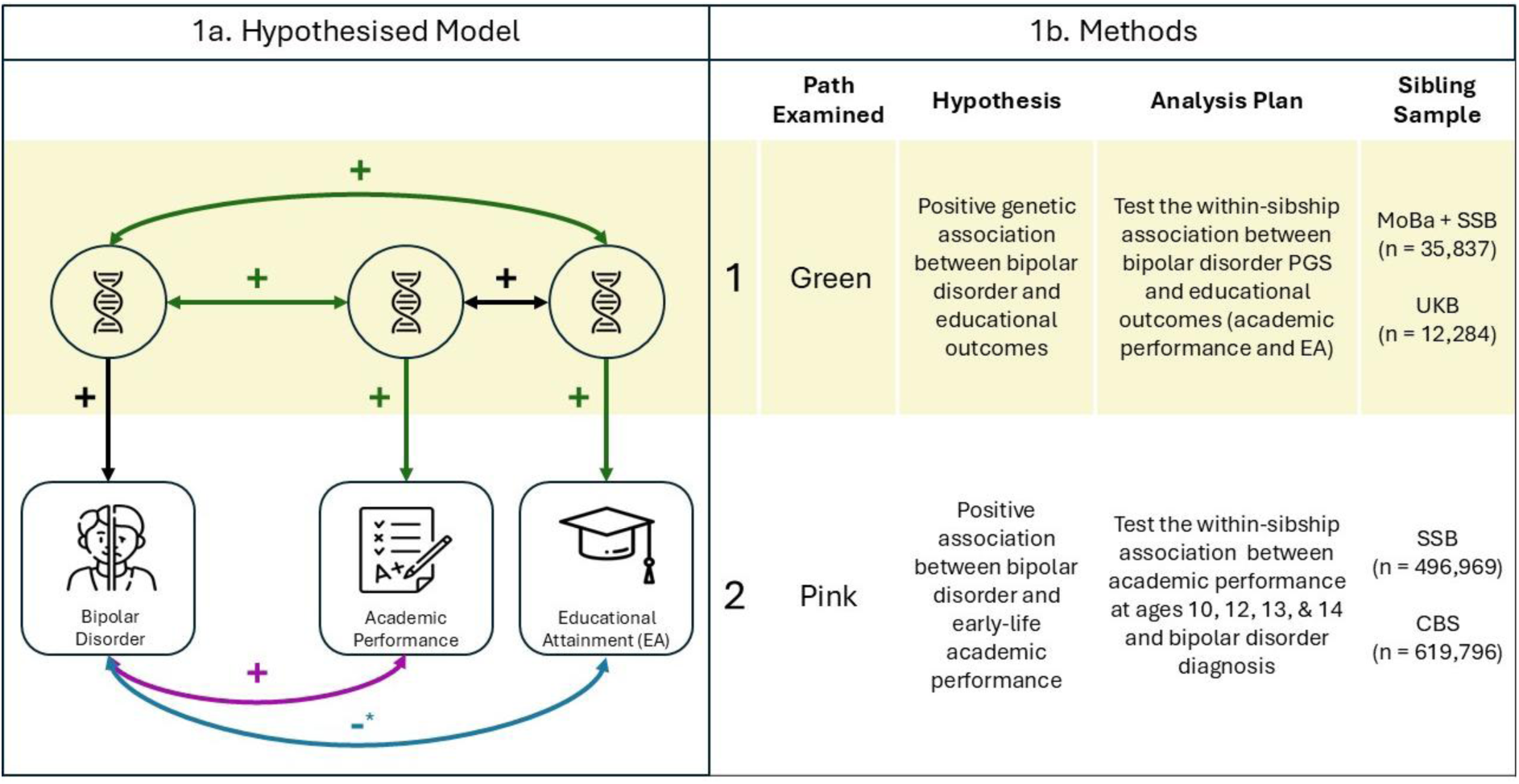
Summary of study hypotheses and methods. 1a represents the hypothesised model of the relationship between bipolar disorder and educational outcomes. The squares represent measured traits and the circles represent genetic predispositions for each trait. Hypothesis 1 is represented by the green pathway, which posits a positive genetic relationship between bipolar disorder and educational outcomes (educational attainment (EA) academic performance). Hypothesis 2 is represented by the pink pathway, which posits a positive relationship between bipolar disorder and early-life academic performance. The blue pathway represents the association between bipolar disorder and long-term educational outcomes such as EA. In our study, we tested the green and pink pathways using the methods outlined in 1b. *the negative association between bipolar disorder and EA (blue pathway) has been reported previously.^6,28^

## Methods

Ethics approval was granted by the Scientific and Ethical Review Board (VCWE) of the Faculty of Behavioural and Movement Sciences, Vrije Universiteit Amsterdam (VCWE-2024-127). Ethical approvals for individual data sources are provided in **Supplementary Methods 1**. This study was pre-registered on the Open Science Framework platform (https://osf.io/tpmj5). Deviations to the pre-registration are outlined in **Supplementary Methods 2**.

### Study Design

We used two complementary analyses, first testing the association between bipolar disorder PGS and educational outcomes (EA and academic performance), and secondly the association between academic performance and subsequent bipolar disorder diagnosis. We employed a sibling comparison design, which assumes that siblings are matched for many familial and socioeconomic factors, as well as ∼50% of their DNA. This facilitates a natural experiment using observational data wherein siblings form a well-matched case-control study.^29,30^ The sibling comparison regression model consists of two components; the between-sibship estimate, which captures differences across families and the within-sibship estimate, which captures associations independent of between-family confounding. We also performed all analyses using population-based regressions to contrast against the within-sibship estimates. Data cleaning procedures for each cohort are outlined in **Supplementary Methods 3**.

### Study Populations

#### I. The Norwegian Mother, Father and Child Cohort Study (MoBa)

MoBa is a population-based pregnancy cohort, which includes approximately 114,500 children, 95,200 mothers, and 75,200 fathers recruited between 1999 and 2008. Women consented to participation in 41% of the pregnancies. Questionnaire data and blood samples were obtained from parents and children during pregnancy and at birth.^31,32^ The pipeline used for quality-control, phasing, and imputation of genotype data in MoBa is described elsewhere.^33,34^

#### II. UK Biobank (UKB)

UKB is a large-scale prospective cohort study with extensive phenotype and genotype data collected from about 500,000 individuals in the UK, aged between 40 and 69 at the time of recruitment. Participants were recruited across diverse socioeconomic and ethnic backgrounds as well as a balance of urban and rural settings.^35,36^ Genotype data quality control procedures for UKB are describe elsewhere and in **Supplementary Methods 4**.^36^

#### III. Statistics Norway (SSB)

SSB maintains the Norwegian population registry, providing extensive data for research and administrative purposes. The National Education Database (NuDB) provides academic data, available for the years 2007 to 2025. This was linked to diagnostic data from the Norwegian Control and Payment of Health Reimbursements Database (KUHR), which provides information on bills reimbursed by the State from primary and specialised health care institutions between 2006 and 2025.

#### IV. Statistics Netherlands (CBS)

The Dutch national registry maintained by CBS provides data on a wide range of topics including education, career, social life, and healthcare. Academic performance data were available for 2006–2014, and diagnostic data for 2011–2021. However, diagnoses beyond 2016 were provided under a broader label “bipolar and other mood disorders”, which doesn’t distinguish between bipolar disorder cases from other substance-induced mood disorders. So we restricted analyses to the dataset with detailed diagnostic labels ending in 2016 (**Supplementary Methods 5**). The extended diagnostic data (through 2021) was used for a supplementary analysis. Diagnoses were based on the care trajectories of people getting specialized mental health care in the Netherlands.

### Measures

#### I. Bipolar Disorder Polygenic Scores (PGS)

In UKB and MoBa, bipolar disorder PGS were constructed using LDPred2-auto,^37^ a Bayesian model that estimates the SNP effect sizes from GWAS summary statistics while accounting for linkage disequilibrium, using assumptions about the trait’s polygenicity and SNP heritability (**Supplementary Methods 5**). Summary statistics were obtained from the most recent large-scale GWAS of bipolar disorder.^38^ The HapMap3+ LD Reference Panel was used to estimate posterior SNP effect sizes, which were multiplied by individual genotype dosages and summed to generate PGS.^39^

#### II. Educational Attainment (EA)

The highest educational qualification was indexed using The Norwegian Standard Classification of Education (NUS2000) codes in MoBa and the variable ‘Qualifications’ (UKB ID: 6138) in UKB. The Norwegian and UKB educational codes were first transformed into the corresponding International Standard Classification of Education (ISCED 2011) codes and subsequently converted into years of education (**Supplementary Methods 5**). We included individuals who were 30 years old or older at the time of measurement, to allow sufficient time to attain their maximum EA.

#### III. Academic Performance

From SSB, we obtained national test scores for mathematics and reading comprehension (in Norwegian) at ages 10, 13, and 14 and for English at ages 10 and 13. Additionally, we calculated a total score as the mean of mathematics and reading comprehension, excluding English to ensure comparability across years. In CBS, we extracted national test scores for mathematics, reading comprehension (in Dutch), language (Dutch), and total score measured at age 12.

#### IV. Bipolar Disorder Diagnosis

In SSB, individuals were considered as cases if they had at least five recorded diagnoses for bipolar disorder from any primary or specialized healthcare institutions. This threshold was established to exclude those with provisional or transient diagnoses. In CBS, diagnostic information was derived from only specialized mental health care services, and so individuals with just one registered diagnosis were considered as a case. In both cohorts, those without a diagnosis were considered as controls. We selected individuals who were at least 15 years old by the final year of available diagnostic data, which was 2025 in SSB and 2016 in CBS.

### Statistical Analysis

Across all models, we account for familial clustering by adjusting standard errors using cluster-robust variance estimators using the R packages sandwich and lmtest.^40,41^ Sex, birth year, and birth order were added as covariates in all analyses. Continuous variables were standardised to enable comparability and replication.

#### I. Analysis 1: Genetic association of bipolar disorder PGS and educational outcomes

Using two separate models, we modelled educational outcomes (EA and academic performance) as the outcome and bipolar disorder PGS as the predictor based on preregistered simulations which indicated greater statistical power compared to the alternative of using EA PGS to predict bipolar disorder diagnosis (**Supplementary Methods 6**). We regressed the educational outcome of each sibling on the mean value of the family’s bipolar disorder PGS (between-sibship effect) and the deviation of the sibling’s bipolar disorder PGS from their family mean (within-sibship effect). The formula for the sibling comparison regression model was as follows.

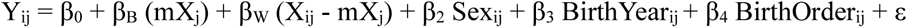

Where Y_ij_ = EA of individual i in family j,

β_B_ = between-sibship effect,

mX_j_ = mean value of the bipolar disorder PGS for the family j,

β_W_ = within-sibship effect,

X_ij_ = bipolar disorder PGS of individual i in family j

For this model, the hypothesised positive association between bipolar disorder PGS and long-term EA may be attenuated among individuals who develop the disorder, as illness onset can disrupt educational trajectories. Therefore, we also ran the main EA model with bipolar disorder diagnosis included as a covariate, to examine whether the association changed after correcting for diagnostic status.

#### II. Analysis 2: Phenotypic association of academic performance and bipolar disorder diagnosis

Cox proportional hazards regression was used to account for those who remained undiagnosed by the end of the observation period. Undiagnosed individuals were right-censored using the age at their last available diagnostic record (2025 in SSB and 2016 in CBS). The formula for the sibling comparison regression model using Cox proportional hazards regression was as follows.

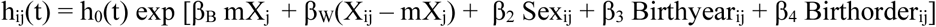

where h_ij_(t) = hazard of bipolar disorder diagnosis at time t for individual i in family j,

h_0_(t) = unspecified baseline hazard,

β_B_ = between-family effect,

mX_j_ = mean value of educational achievement of family j,

β_W_ = within-sibship effect,

X_ij_ = educational achievement of individual i in family j

As an exploratory analysis, we ran the models for ages 13 and 14 with scores from previous ages included in the regression model as covariates. This adjusted model specifies how the relative change in academic performance across ages is related to bipolar disorder diagnosis, above and beyond an individual’s previous academic performance.

## Results

Descriptives from each analysis are provided in **Table 1** (Full descriptives in **Supplementary Table 5**).

**Table 1.** Sample Characteristics. Note: MoBa = The Norwegian Mother, Father and Child Cohort; SSB = Statistics Norway; UKB = UK Biobank; CBS = Statistics Netherlands; M = Mean; SD = Standard Deviation; PGS = Polygenic Scores; EA = Educational Attainment; M = Mean; SD = Standard Deviation. Data from SSB was used in linkage with the Norwegian Control and Payment of Health Reimbursements Database (KUHR). The sibling samples were used to estimate between- and within-sibship associations.

| Variable | Analysis 1 |  |  |  |  |  | Analysis 2 |  |  |  |
|---|---|---|---|---|---|---|---|---|---|---|
|  | Bipolar PGS and EA |  |  |  | Bipolar PGS and academic performance |  | Academic performance and bipolar disorder diagnosis |  |  |  |
|  | MoBa + SSB |  | UKB |  | MoBa + SSB |  | SSB |  | CBS |  |
|  | **Population Sample** | **Sibling Sample** | **Population Sample** | **Sibling Sample** | **Population Sample** | **Sibling Sample** | **Population Sample** | **Sibling Sample** | **Population Sample** | **Sibling Sample** |
| Sample Size, N | 121,550 | 16,746 | 192,726 | 12,284 | 74,104 | 19,091 | 1,026,024 | 496,969 | 1,144,287 | 619,796 |
| No. of Families, N | 110,507 | 8,107 | 184,667 | 5,997 | 64,290 | 9,277 | 626,043 | 208,350 | 807,073 | 286,658 |
| Birth Year, Range | 1960 - 1992 | 1960 – 1990 | 1934 - 1970 | 1937 - 1970 | 1999-2009 | 1999-2009 | 1993 - 2010 | 1993 - 2010 | 1992 - 2001 | 1992 - 2001 |
| Females, n(%) | 72,973 (60.04%) | 10,077 (60.18%) | 103,874 (53.9%) | 7139 (58.1%) | 36,368 (48.08%) | 9,354 (49%) | 502,113 (48.94%) | 242,262 (48.75%) | 572070 (49.99%) | 309227 (49.89%) |
| Bipolar Disorder Cases, n(%) | 1375 (1.13%) | 191 (1.14%) | .. | .. | .. | .. | 4995 (0.49%) | 2654 (0.53%) | 378 (0.03%) | 173 (0.03%) |
| Age at Diagnosis, M(SD) | 38.24 (7.22) | 38.71 (6.8) | .. | .. | .. | .. | 21.3 (3.83) | 21.21 (3.71) | 18.7 (2.56) | 18.59 (2.49) |
| EA, M(SD) | 16.79 (3.39) | 17.16 (3.23) | 15.42 (4.94) | 15.08 (5) | .. | .. | .. | .. | .. | .. |
| Bipolar Disorder PGS, M(SD) | 2.76 (0.6) | 2.76 (0.6) | 3.06 (2.6) | 3.03 (2.62) | 2.76 (0.6) | 2.76 (0.6) | .. | .. | .. | .. |

### Analysis 1: Bipolar disorder PGS and educational outcomes

The within-sibship association between bipolar disorder PGS and EA was positive and significant in MoBa (β_W_ = 0.034, 95% CI [0.012, 0.055]), indicating an increase of 0.22 years in EA per SD increase in bipolar disorder PGS. The within-sibship association in UKB was also positive but not significant (**Figure 2a** and **Supplementary Table 6**).

We examined whether adjustment for bipolar disorder diagnosis altered the association between bipolar disorder PGS and EA to remove any potential influence of bipolar disorder symptoms on EA; all the significant associations from our main analyses remained when doing so (adjusted estimates in **Figure 2a** and **Supplementary Results 7**).

We also examined associations between bipolar disorder PGS and academic performance at ages 10, 13, and 14. Bipolar disorder PGS was positively associated with academic performance at age 10 for total score and reading, but not for mathematics. These associations were no longer significant at later ages, except for reading scores at age 14 (**Figure 2b** and **Supplementary Table 8**).

**Figure 2.**
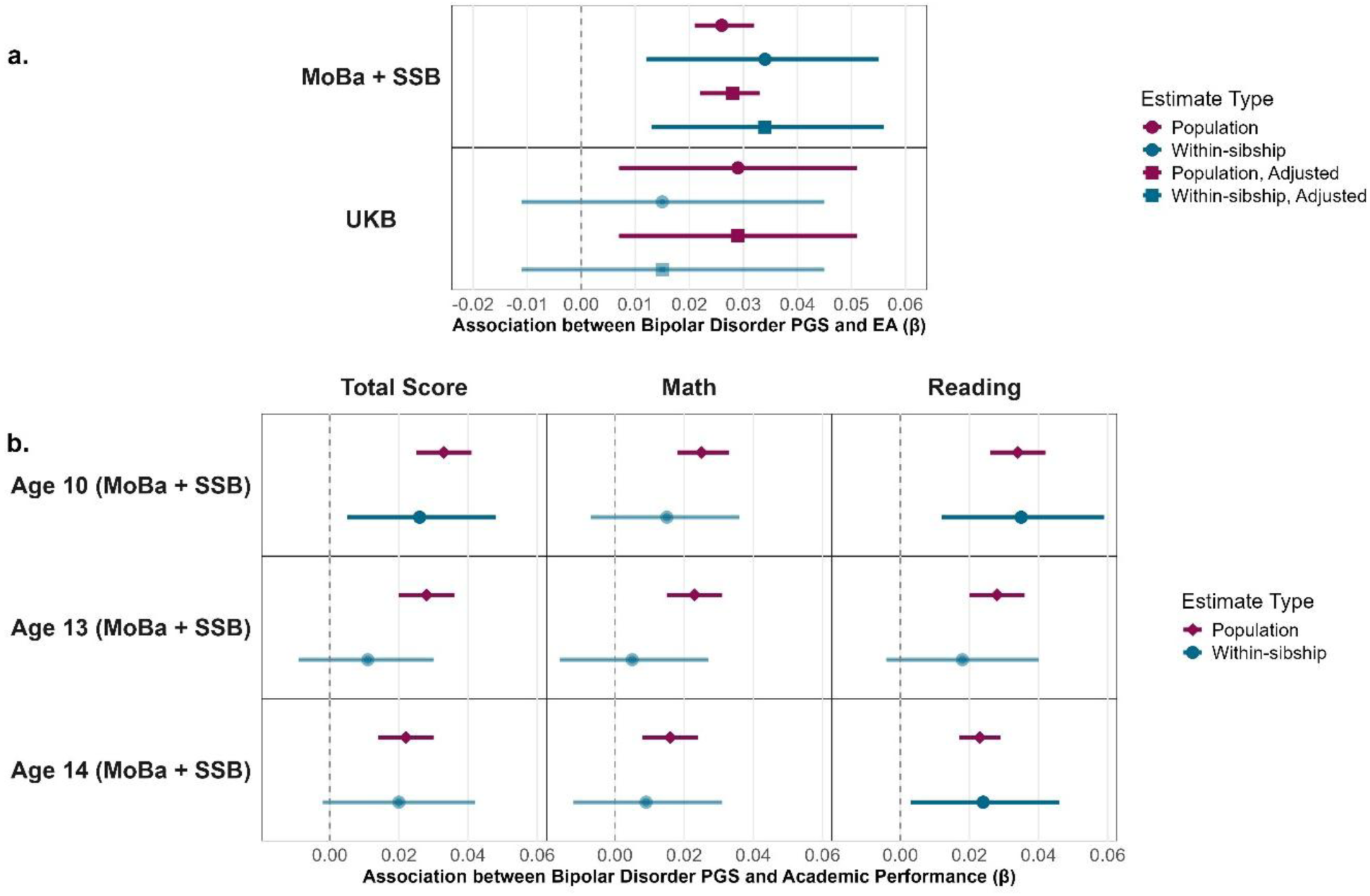
Associations between bipolar disorder PGS and educational outcomes – educational attainment (EA) and academic performance at ages 10, 13, 14 in the cohorts MoBa + SSB and UKB. Pink represents the population estimate and blue represents the within-sibship estimate obtained from the sibling sample. The lines represent 95% confidence intervals. 2a presents the associations between bipolar disorder PGS and EA. Adjusted estimates for EA analysis are corrected for bipolar disorder diagnosis. The population estimates for UKB are represented by the between-family estimates. 2b presents the associations between bipolar disorder PGS and academic performance at ages 10, 13, and 14. Results for English test scores are provided in Supplementary Figure 1. Reading comprehension test was administered in Norwegian.

### Analysis 2: Academic performance and bipolar disorder diagnosis

Individuals later diagnosed with bipolar disorder had higher total scores than their siblings at age 10, as shown by the hazards ratio (HR) greater than 1 (HR_W_ = 1.11, 95% CI [1.02, 1.22]). This positive association between academic performance and bipolar disorder diagnosis was also reflected in reading scores, with high scores corresponding to a 17% higher hazard (HR_W_ = 1.17, 95% CI [1.07, 1.28]) of obtaining a bipolar disorder diagnosis respectively per unit-SD increase in test score (**Figure 3** and **Supplementary Table 9**). The hazard percentage is calculated as (HR – 1) * 100.

Similarly, using test scores from the Netherlands (CBS) at age 12, higher total score was associated with a significantly higher hazard of bipolar disorder diagnosis (HR_W_ = 1.59, 95% CI [1.16, 2.19]). This positive association was found across all individual test scores namely mathematics, reading comprehension, and language.

At age 13, this positive association was no longer observed in SSB, as the HR was not found to be significantly different than 1. At age 14, the associations remained null and even turned negative for mathematics scores; the sibling with a diagnosis of bipolar disorder performed worse than their unaffected sibling at age 14, as indicated by the HR value lower than 1 (HR_W_ = 0.90, 95% CI [0.83, 0.98]).

As an exploratory analysis, we examined the association between the relative change in academic performance and subsequent bipolar disorder diagnosis by adjusting for performance at previous ages (adjusted estimates in **Figure 3** and **Supplementary Table 10**). Correcting for the scores from age 10, the association in age 13 turned negative such that the sibling with bipolar disorder performed worse compared to their unaffected sibling (HR_W_ = 0.84, 95% CI [0.73, 0.96]), corresponding to a 19% higher hazards of diagnosis for a unit-SD decline in academic performance (calculated as ((1/HR) – 1 * 100) for HR less than 1). This trend was also observed for mathematics scores (HR_W_ = 0.88, 95% CI [0.78, 0.99]).

Correcting for scores from age 10 and 13, the association between the decline in academic performance and bipolar disorder diagnosis became even stronger at age 14 (HR_W_ = 0.72, 95% CI [0.61, 0.85]), corresponding to a 38% higher hazard of diagnosis per unit-SD decline in test score. This was also reflected in mathematics (HR_W_ = 0.78, 95% CI [0.67, 0.90]) as well as reading scores (HR_W_ = 0.86, 95% CI [0.75, 0.98]).

Given that our primary analyses assumed linear associations, we next tested whether the relationship between academic performance and bipolar disorder diagnosis deviated from linearity; we found no strong evidence of non-linearity, except for mathematics and English scores at age 10, for which the spline model had a significant better fit than a linear model (**Supplementary Results 8**).

Next, we considered whether the wide confidence intervals observed in our CBS estimates reflected low statistical power due to the small number of bipolar disorder cases in this sample. To address this, we repeated our analyses using the extended dataset available through 2021, which offered a larger number of cases but less precise case ascertainment as it doesn’t distinguish between bipolar disorder diagnoses from other substance-induced mood disorders. This yielded population-level estimates similar to the primary analysis in direction and magnitude across all tests, but the within-family associations were not significant (**Supplementary Results 9**).

**Figure 3.**
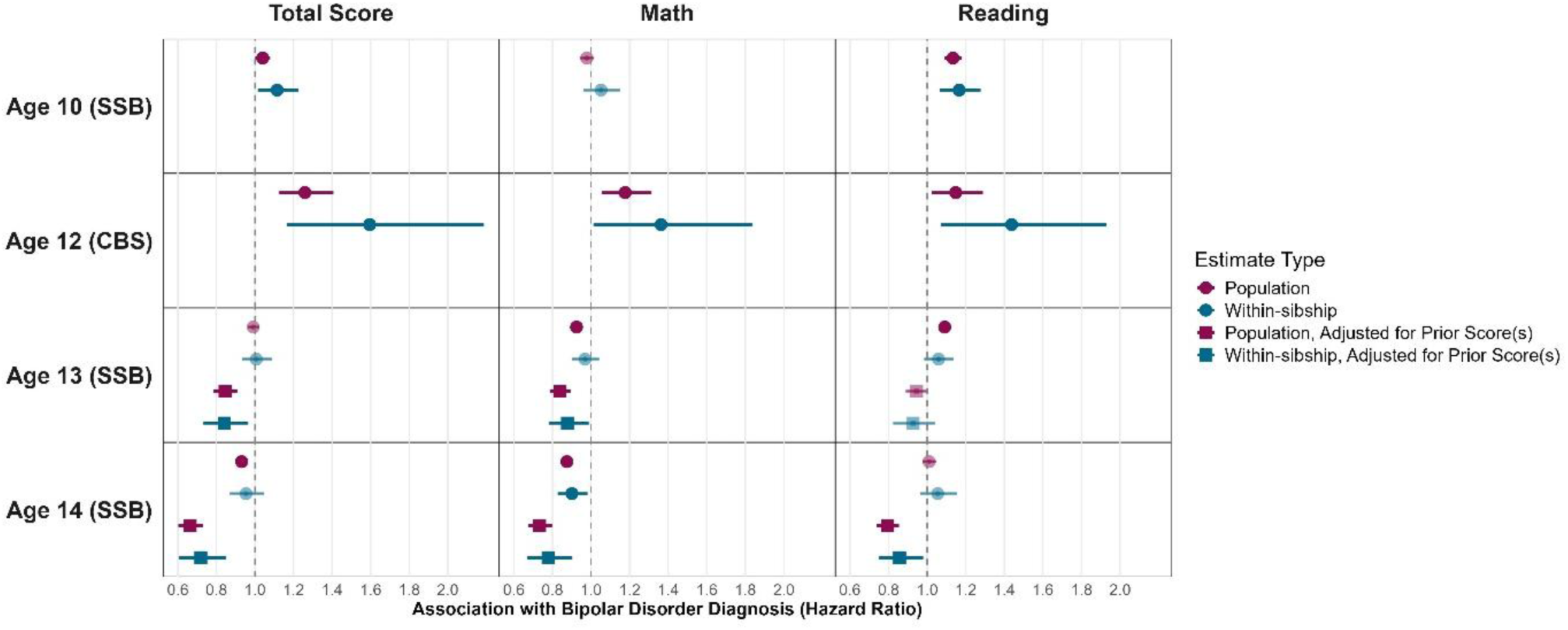
Associations between academic performance at ages 10, 12, 13, 14 and bipolar disorder diagnosis in the cohorts SSB and CBS. The estimates at ages 10, 13, and 14 are from SSB and the estimates at age 12 are from CBS. Reading comprehension tests are administered in Norwegian in SSB and Dutch in SSB. The figure including English scores in SSB and language scores in CBS is provided in Supplementary Figure 2. Pink represents the population estimate and blue represents the within-sibship estimate obtained from the sibling sample. The lines represent 95% confidence intervals; these are not necessarily symmetric around the estimate as hazard ratios from Cox regression are estimated on the log scale and then exponentiated.

## Discussion

In the current study, we used a sibling comparison design across four cohorts to examine the complex association between bipolar disorder and educational outcomes. We found that higher probabilistic genetic risk for bipolar disorder, indexed using PGS, was associated with higher EA, as well as higher early-life academic performance at age 10 but not at ages 13 and 14. Moreover, using observed academic performance and diagnostic data, we found that individuals with a diagnosis of bipolar disorder academically outperformed their unaffected sibling at ages 10 and 12. However, this academic advantage diminished by ages 13 and 14, with a decline in academic performance significantly related to bipolar disorder. Our findings are consistent with the suggested model which posits that bipolar disorder and educational outcomes share a positive genetic association, reflected in early academic benefit, but that the emergence of bipolar disorder symptoms disrupts educational trajectories, ultimately resulting in poor long-term educational outcomes. Taken together, we discuss how our research can help explain conflicting findings in literature, provide epidemiological and clinically-relevant insights as well as highlight potential avenues for future research.

Bipolar disorder PGS was positively associated with EA, which corroborates previous observational findings.^12–15^ In MoBa, this association persisted even after accounting for shared familial confounding at the within-sibship level. In UKB, the positive association observed at the population level was no longer significant at the within-sibship level, consistent with prior work^42^ This discrepancy could reflect inter-societal variation in educational systems, differences in phenotypic measurement, or even selection into the two cohorts; UKB is skewed toward older, healthier adults, whereas MoBa is a pregnancy cohort typically including younger participants.^43^

In addition to EA, we tested the association between bipolar disorder PGS and early-life academic performance in MoBa. While we found significant positive associations at age 10, they were no longer significant at ages 13 and 14, suggesting that genetic associations are not consistent across development. This may reflect exogenous influences on education emerging in early adolescence, such as the transition from primary to lower secondary school in Norway at age 13, bringing about changes in schooling environment and peer groups.^44^ The divergence with EA also implies that academic performance and EA are not equivalent constructs; national test scores resemble cognitive tests and are less influenced by socioeconomic influences.^45^

Our phenotypic investigation of academic performance across ages 10 to 14 revealed that those with bipolar disorder perform well in school at ages 10 and 12 compared to their unaffected siblings, indicated by a 11 to 59% higher hazard of receiving a diagnosis per unit-SD increase in test score. This academic advantage attenuated by ages 13 and 14, with the associations no longer significant.

Extending this longitudinal analysis to an adjusted model, we controlled for the influence of previous test scores to reveal that a decrease in academic performance at ages 13 and 14 is associated with bipolar disorder diagnosis; a unit-SD decline in performance at age 13 was associated with 8 - 19% increased hazard of getting a diagnosis, and a decline at age 14 was associated with 17 - 39% increased hazard. The emergence of signs of bipolar disorder at such an early age is consistent with previous evidence of a prolonged prodromal period, such as depressive symptoms emerging in mid-adolescence, several years before the first (hypo)manic episode and diagnosis.^47,48^ Additionally, our findings suggest that relative changes in functional performance may be a better marker of psychopathology risk than absolute academic standing, highlighting the importance of longitudinal data in research as well as continuous observations in clinical practice to get a comprehensive understanding of psychiatric epidemiology.

Comparison with previous conflicting evidence is complicated due to key differences in educational and healthcare systems (Supplementary Methods 5), as well as methodological, and analytical approaches across cohorts.^22–25^ For example, national test scores in Norway are mainly used to monitor school progress, whereas in the Netherlands, they determine selection into secondary school tracks and therefore involve preparation and pressure. Moreover, our analyses revealed age-specific effects of bipolar disorder on academic performance across the developmental trajectory, whereas previous studies with conflicting findings examine academic performance measured at a single age, which limits insight into the academic progression of those with bipolar disorder within each cohort.^22–25^

The magnitude of associations differed across the type of tests; language/English and reading comprehension showed mostly positive associations with subsequent bipolar disorder, whereas mathematics followed a different developmental pattern, with little evidence of an early advantage but the most pronounced decline during adolescence. This developmental pattern aligns with previous evidence of poor arithmetic skills among those with bipolar disorder but higher linguistic ability and creativity at the genetic as well as phenotypic level._2_^5,49,50^

Our findings should be interpreted in light of some limitations. First, UKB and MoBa are among the largest genotyped cohorts currently available, but are subject to sampling bias, with participants generally being healthier and more highly educated than the general population.^51–53^ Future studies should undertake collaborative efforts to meta-analyse results from diverse genotyped cohorts.

Second, while the sibling design accounts for unmeasured confounding factors shared by siblings, it does not control for non-shared confounding and measurement error, which has been shown to affect within-sibship estimates.^54^ Measurement error in the exposure attenuates within-sibship estimates more than population estimates, particularly when siblings’ exposure values are correlated, relevant for our PGS analyses.^55^

Thirdly, the period of follow-up for bipolar disorder diagnosis was limited to approximately age 30 in SSB and age 24 in CBS. To address this limitation, we employed a Cox proportional hazards model to account for individuals who remain undiagnosed by the end of the observation period. But other issues like cross-cohort discrepancies in case definitions, misdiagnosis and undetected illness remain, potentially attenuating the associations studied.^56^

Finally, while we know from previous literature and the current study that high academic performance is a key feature of bipolar disorder,^20,57^ it remains unclear whether the decline is specific to bipolar disorder. This warrants comparison with longitudinal academic trajectories of those with other severe mental health issues. In addition, our findings do not provide insight into the specific processes associated with the academic decline in mid-adolescence, and may be related to depressive symptoms, non-specific mental health difficulties, cognitive decline, or other non-cognitive factors.^45,58,59^ Investigation into these specific vulnerabilities underlying the educational disruption can provide measurable indicators of emerging bipolar disorder with substantial clinical and interventional potential.

## Conclusion

Using two complementary genetically-informed approaches across multiple large-scale cohorts and national registry datasets, we find evidence of a positive genetic association between bipolar disorder and educational outcomes. Individuals with bipolar disorder outperformed their unaffected sibling in academic performance at ages 10 and 12. However, those with bipolar disorder diagnoses experienced a decline in their performance at ages 13 and 14, providing new epidemiological and clinically-relevant insights.

## Data Availability

Access to data used in this research is subject to specific application procedures for each cohort. More information can be found at https://www.ukbiobank.ac.uk/ for UKB, https://www.ssb.no/en/data-til-forskning/utlan-av-data-til-forskere for SSB, https://www.fhi.no/en/studies/moba/for-forskere-artikler/research-and-data-access/ for MoBa, and and https://www.cbs.nl/en-gb/our-services/customised-services-microdata/microdata-conducting-your-own-research for CBS.

## Code and Data Availability

All code used for data analyses is available at https://github.com/VaishnaviMadhavan/BipolarDisorder-EA.

## Contributions

Conceptualisation, study design, methodology: VKM, WAA, PAD, MN. Resources, Data curation, Funding acquisition: WAA, VKM, EY. Data analysis: VKM, WAA, PAD, MN. Supervision: WAA, PAD, MN, MB, JJH. Writing original draft: VKM, WAA, PAD. Writing review and editing: VKM, WAA, PAD, MN, MB, JJH, ZA, EY. The first author VKM had access to all datasets used in the current study. WAA had access to UKB and CBS data and PAD had access to SSB and MoBa data. All authors contributed to and approved the final version of the manuscript.

## Acknowledgements

The authors would like to extend sincere thanks for the contributions of SUBPU data managers Clara Timpe and Oda van Jole, as well as Chris Rayner for assistance with making the polygenic scores in MoBa.

VKM and WAA are supported by a Talent and Early Development Grant from the Dutch Ministry of Education, Culture and Sciences (OCW). PAD and EY are supported by the European Union Grant agreement No. 101045526. EY is also supported by the European Union (101073237), the Research Council of Norway (336078, 288083, 331640), and the Swedish Research Council (2024-06499_VR).

The Norwegian registry and MoBa data were used via the project SUBPU, which has agreements with the MoBa and Statistics Norway for data linkage and usage. SUBPU received ethical approval from the Regional Committees for Medical and Health Research Ethics (reference: 2017/2205). The University of Oslo is responsible for the data handling in SUBPU and has conducted a Data Protection Impact Assessment (DPIA) in collaboration with the Norwegian Agency for Shared Services in Education and Research (Sikt; ref. 962088). The data access and management of SUBPU is financed by the Research Council of Norway (RCN), the European Research Council (ERC), and the Department of Psychology at the University of Oslo. All MoBa participants provided informed consent and data were processed using procedures compliant with national data protection regulations. MoBa is regulated by the Norwegian Health Registry Act.

The data management, storage, and analyses for Norwegian data were performed on the secure data “Tjeneste for Sensitive Data” (TSD) facilities, owned by the University of Oslo, operated and developed by the TSD service group at the University of Oslo, IT-Department (USIT). Resources provided by Sigma2, the National Infrastructure for High Performance Computing and Data Storage in Norway (UNINETT), was used for analyses (#NS9867S).

The Norwegian Mother, Father and Child Cohort Study is supported by the Norwegian Ministry of Health and Care Services and the Ministry of Education and Research. We are grateful to all the participating families in Norway who take part in this on-going cohort study.

This research was conducted in part using ODISSEI, the Open Data Infrastructure for Social Science and Economic Innovations (https://ror.org/03m8v6t10) Access to Statistics Netherlands (CBS) was facilitated by funding from the Microdata Access Grant (MAG) ODISSEI under the project ‘Bipolar Disorder and Educational Attainment: Deciphering a Complex Relationship using Genetic and Sibling Differences’ (project number 3157).

UK Biobank (UKB) Data used in this research was accessed under the application number 531743. UKB data are available to bona fide researchers upon approval of a research project. Further information can be found at: https://www.ukbiobank.ac.uk/.

## References

1 Fajutrao L, Locklear J, Priaulx J, Heyes A. A systematic review of the evidence of the burden of bipolar disorder in Europe. Clin Pract Epidemiol Ment Health 2009; 5: 3.

2 Marwaha S, Durrani A, Singh S. Employment outcomes in people with bipolar disorder: a systematic review. Acta Psychiatrica Scandinavica 2013; 128: 179–93.

3 Wyatt RJ, Henter I. An economic evaluation of manic-depressive illness–1991. Soc Psychiatry Psychiatr Epidemiol 1995; 30: 213–9.

4 Mojtabai R, Stuart EA, Hwang I, Eaton WW, Sampson N, Kessler RC. Long-Term Effects of Mental disorders on Educational Attainment In the National Comorbidity Survey Ten-Year Follow-up. Soc Psychiatry Psychiatr Epidemiol 2015; 50: 1577–91.

5 Tempelaar WM, Termorshuizen F, MacCabe JH, Boks MPM, Kahn RS. Educational achievement in psychiatric patients and their siblings: a register-based study in 30 000 individuals in The Netherlands. Psychological Medicine 2017; 47: 776–84.

6 Demange PA, Boomsma DI, van Bergen E, Nivard MG. Educational attainment and psychiatric diagnoses: a national registry data and two-sample Mendelian randomization study. Nat Mental Health 2024; 2: 668– 79.

7 Adams CD. A multivariable Mendelian randomization to appraise the pleiotropy between intelligence, education, and bipolar disorder in relation to schizophrenia. Sci Rep 2020; 10: 6018.

8 Baranova A, Cao H, Zhang F. Exploring the influences of education, intelligence and income on mental disorders. General Psychiatry 2024; 37: e101080.

9 Bulik-Sullivan B, Finucane HK, Anttila V, et al. An Atlas of Genetic Correlations across Human Diseases and Traits. Nat Genet 2015; 47: 1236–41.

10 Shang M-Y, Wu Y, Zhang C-Y, et al. Bidirectional genetic overlap between bipolar disorder and intelligence. BMC Medicine 2022; 20: 464.

11 Yuan S, Xiong Y, Michaëlsson M, Michaëlsson K, Larsson SC. Genetically predicted education attainment in relation to somatic and mental health. Sci Rep 2021; 11: 4296.

12 Hagenaars SP, Harris SE, Davies G, et al. Shared genetic aetiology between cognitive functions and physical and mental health in UK Biobank (N=112 151) and 24 GWAS consortia. Mol Psychiatry 2016; 21: 1624–32.

13 Hazak A, Liuhanen J, Kantojärvi K, et al. Polygenic Risks for Mood Disorders and Economic Well-being: Study of Finnish Cohorts. Depression and Anxiety 2026; 2026: 1008569.

14 Mistry S, Harrison JR, Smith DJ, Escott-Price V, Zammit S. The use of polygenic risk scores to identify phenotypes associated with genetic risk of bipolar disorder and depression: A systematic review. Journal of Affective Disorders 2018; 234: 148–55.

15 Wu C-S, Hsu C-L, Lin M-C, et al. Association of polygenic liabilities for schizophrenia and bipolar disorder with educational attainment and cognitive aging. Transl Psychiatry 2024; 14: 472.

16 Choi SW, Mak TS-H, O’Reilly PF. Tutorial: a guide to performing polygenic risk score analyses. Nat Protoc 2020; 15: 2759–72.

17 Gale CR, Batty GD, McIntosh AM, Porteous DJ, Deary IJ, Rasmussen F. Is bipolar disorder more common in highly intelligent people? A cohort study of a million men. Mol Psychiatry 2013; 18: 190–4.

18 Koenen KC, Moffitt TE, Roberts AL, et al. Childhood IQ and adult mental disorders: a test of the cognitive reserve hypothesis. Am J Psychiatry 2009; 166: 50–7.

19 Tiihonen J, Haukka J, Henriksson M, et al. Premorbid intellectual functioning in bipolar disorder and schizophrenia: results from a cohort study of male conscripts. Am J Psychiatry 2005; 162: 1904–10.

20 Vreeker A, Boks MPM, Abramovic L, et al. High educational performance is a distinctive feature of bipolar disorder: a study on cognition in bipolar disorder, schizophrenia patients, relatives and controls. Psychol Med 2016; 46: 807–18.

21 Smith DJ, Anderson J, Zammit S, Meyer TD, Pell JP, Mackay D. Childhood IQ and risk of bipolar disorder in adulthood: prospective birth cohort study. BJPsych Open 2015; 1: 74–80.

22 MacCabe JH, Lambe MP, Cnattingius S, et al. Excellent school performance at age 16 and risk of adult bipolar disorder: national cohort study. Br J Psychiatry 2010; 196: 109–15.

23 Gyllenberg D, Ristikari T, Kelleher I, Kääriälä A, Gissler M. School performance and later diagnoses of nonaffective psychoses, bipolar disorder, and depression. Acta Psychiatrica Scandinavica 2022; 146: 420– 9.

24 Kendler KS, Ohlsson H, Mezuk B, Sundquist K, Sundquist J. A Swedish National Prospective and Co-relative Study of School Achievement at Age 16, and Risk for Schizophrenia, Other Nonaffective Psychosis, and Bipolar Illness. Schizophrenia Bulletin 2016; 42: 77–86.

25 Pedersen SD, Østergaard SD, Petersen L. The association between school exam grades and subsequent development of bipolar disorder. Acta Neuropsychiatrica 2018; 30: 209–17.

26 Tsuchiya KJ, Agerbo E, Byrne M, Mortensen PB. Higher socio-economic status of parents may increase risk for bipolar disorder in the offspring. Psychol Med 2004; 34: 787–93.

27 Rowland TA, Marwaha S. Epidemiology and risk factors for bipolar disorder. Therapeutic Advances in Psychopharmacology 2018; 8: 251–69.

28 Hakulinen C, Musliner KL, Agerbo E. Bipolar disorder and depression in early adulthood and long-term employment, income, and educational attainment: A nationwide cohort study of 2,390,127 individuals. Depress Anxiety 2019; 36: 1080–8.

29 Donovan SJ, Susser E. Commentary: Advent of sibling designs. Int J Epidemiol 2011; 40: 345–9.

30 Sjölander A, Zetterqvist J. Confounders, Mediators, or Colliders: What Types of Shared Covariates Does a Sibling Comparison Design Control For? Epidemiology 2017; 28: 540–7.

31 Paltiel L, Anita H, Skjerden T, et al. The biobank of the Norwegian Mother and Child Cohort Study – present status. Norsk Epidemiologi 2014; 24. DOI:10.5324/nje.v24i1-2.1755.

32 Brandlistuen RE, Kristjansson D, Alsaker E, et al. Cohort Profile Update: The Norwegian Mother, Father and Child Cohort (MoBa). Int J Epidemiol 2025; 54: dyaf139.

33 Corfield EC, Frei O, Shadrin AA, et al. The Norwegian Mother, Father, and Child cohort study (MoBa) genotyping data resource: MoBaPsychGen pipeline v.1. 2022; : 2022.06.23.496289.

34 Corfield EC, Shadrin AA, Frei O, et al. Family genetic designs in MoBa provide insights into health and functioning. Nature 2026; : 1–9.

35 Sudlow C, Gallacher J, Allen N, et al. UK Biobank: An Open Access Resource for Identifying the Causes of a Wide Range of Complex Diseases of Middle and Old Age. PLoS Medicine 2015; 12. DOI:10.1371/journal.pmed.1001779.

36 Bycroft C, Freeman C, Petkova D, et al. The UK Biobank resource with deep phenotyping and genomic data. Nature 2018; 562: 203–9.

37 Privé F, Arbel J, Vilhjálmsson BJ. LDpred2: better, faster, stronger. Bioinformatics 2020; 36: 5424–31.

38 O’Connell KS, Koromina M, van der Veen T, et al. Genomics yields biological and phenotypic insights into bipolar disorder. Nature 2025; 639: 968–75.

39 Privé F, Albiñana C, Arbel J, Pasaniuc B, Vilhjálmsson BJ. Inferring disease architecture and predictive ability with LDpred2-auto. The American Journal of Human Genetics 2023; 110: 2042–55.

40. Zeileis A, Lumley T, Graham N, Koell S. sandwich: Robust Covariance Matrix Estimators. 2026; published online Aug 3. https://cran.r-project.org/web/packages/sandwich/index.html (accessed Aug 11, 2026).

41. CRAN: Package lmtest. https://cran.r-project.org/web/packages/lmtest/index.html (accessed Aug 11, 2026).

42 Weijer MP van de, Demange PA, Pelt DHM, Bartels M, Nivard MG. Disentangling potential causal effects of educational duration on well-being, and mental and physical health outcomes. Psychological Medicine 2024; 54: 1403–18.

43 Fry A, Littlejohns TJ, Sudlow C, et al. Comparison of Sociodemographic and Health-Related Characteristics of UK Biobank Participants With Those of the General Population. Am J Epidemiol 2017; 186: 1026–34.

44 Strand GM. Experiencing the transition to lower secondary school: Students’ voices. International Journal of Educational Research 2019; 97: 13–21.

45 Brookhart SM, Guskey TR, Bowers AJ, et al. A Century of Grading Research: Meaning and Value in the Most Common Educational Measure. Review of Educational Research 2016; 86: 803–48.

46 Berk M, Dodd S, Callaly P, et al. History of illness prior to a diagnosis of bipolar disorder or schizoaffective disorder. J Affect Disord 2007; 103: 181–6.

47 Mesman E, Nolen WA, Reichart CG, Wals M, Hillegers MHJ. The Dutch Bipolar Offspring Study: 12-Year Follow-Up. American Journal of Psychiatry 2013; 170: 542–9.

48 Rajagopal VM, Ganna A, Coleman JRI, et al. Genome-wide association study of school grades identifies genetic overlap between language ability, psychopathology and creativity. Sci Rep 2023; 13: 429.

49 MacCabe JH, Sariaslan A, Almqvist C, Lichtenstein P, Larsson H, Kyaga S. Artistic creativity and risk for schizophrenia, bipolar disorder and unipolar depression: a Swedish population-based case-control study and sib-pair analysis. Br J Psychiatry 2018; 212: 370–6.

50 Schoeler T, Speed D, Porcu E, Pirastu N, Pingault J-B, Kutalik Z. Participation bias in the UK Biobank distorts genetic associations and downstream analyses. Nat Hum Behav 2023; 7: 1216–27.

51 Vejrup K, Magnus P, Magnus M. Lost to follow-up in the Norwegian mother, father and child cohort study. Paediatric and Perinatal Epidemiology 2022; 36: 300–9.

52 Rayner C, Hannigan LJ, Badini I, et al. Quantifying and adjusting for selection biases in the Norwegian Mother, Father and Child Cohort Study using population-wide individual-level registry information. Int J Epidemiol 2026; 55: dyag122.

53 Frisell T, Öberg S, Kuja-Halkola R, Sjölander A. Sibling Comparison Designs: Bias From Non-Shared Confounders and Measurement Error. Epidemiology 2012; 23: 713.

54 Gustavson K, Torvik FA, Davey Smith G, Røysamb E, Eilertsen EM. Familial confounding or measurement error? How to interpret findings from sibling and co-twin control studies. Eur J Epidemiol 2024; 39: 587–603.

55 Persson I, Khamis H. Bias Of The Cox Model Hazard Ratio. J Mod App Stat Meth 2005; 4: 90–9.

56 Bhattacharyya U, John J, Preuss M, Lam M, Lencz T. Cognitive pleiotropy reveals disorder-specific and shared biology for schizophrenia and bipolar disorder. 2026; : 2026.07.07.26357446.

57 Weckström T, Elovainio M, Pulkki-Råback L, et al. School achievement in adolescence and the risk of mental disorders in early adulthood: a Finnish nationwide register study. Mol Psychiatry 2023; 28: 3104– 10.

58 Costa A, Moreira D, Casanova J, et al. Determinants of academic achievement from the middle to secondary school education: A systematic review. Soc Psychol Educ 2024; 27: 3533–72.

