## Supplementary Information for "Bipolar Disorder and Education: Deciphering a Complex Relationship using Genetic and Sibling Differences in a Multi-cohort Study"

**Supplementary Methods 1: Data Sources and Ethical Approvals**

Multiple datasets were used in this study, each subject to specific access procedures and restrictions.
UK Biobank (UKB) Resource was used in this research under the application number 531743. UKB data are available to bona fide researchers upon approval of a research project. Further information can be found at: https://www.ukbiobank.ac.uk/. Ethical approval for UKB was provided by the National Health Service North West Centre for Research Ethics Committee (reference: 11/NW/0382).

Norwegian registry data were obtained from Statistics Norway (SSB; https://www.ssb.no/en) in linkage with the Norwegian Control and Payment of Health Reimbursements Database (KUHR; https://helsedata.no/en/forvaltere/norwegian-directorate-of-health/norwegian-control-and-payment-of-health-reimbursements-database-kuhr/). Access to these data is subject to approval from the Regional Committees for Medical and Health Research Ethics (REC), compliance with the EU General Data Protection Regulation (GDPR), and approval from the data owners. Information and guidelines for accessing microdata from Statistics Norway (SSB) can be found at https://www.ssb.no/en/data-til-forskning/utlan-av-data-til-forskere.

Data from the Norwegian Mother, Father and Child Cohort Study (MoBa; https://www.fhi.no/en/ch/studies/moba/) is handled by the Norwegian Institute of Public Health, and accessible upon application. Instructions for data access are available at: https://www.fhi.no/en/studies/moba/for-forskere-artikler/research-and-data-access/. Ethical approval for MoBa was granted by the Regional Committees for Medical and Health Research Ethics (reference: 2017/2205). Data management is handled by the University of Oslo under agreements with SSB.

Statistics Netherlands (CBS) consists of restricted-access registry microdata which can be accessed for statistical and scientific research. We analysed CBS data under the project 'Bipolar Disorder and Educational Attainment: Deciphering a Complex Relationship using Genetic and Sibling Differences' (project number 3157). Information on access procedures can be obtained via.

**Supplementary Methods 2: Deviations to the Pre-registration**

A few changes were made to the pre-registered analysis plan to improve the rigor and scope of the study.

*Analysis 1*

- Polygenic Score (PGS) Calculation: The PGS were calculated using summary statistics from the most recent and comprehensive GWAS of bipolar disorder (O'Connell et al., 2025), instead of the GWAS originally specified in the pre-registration (Mullins et al., 2021).
- Educational Attainment (EA): For the analysis using Norwegian data, we obtained EA from the National Education Database (NUDB), with the highest possible qualification being a PhD degree. We increased the lower-cut off age for the EA variable from 25 to 30 years of age to allow most individuals to obtain their maximum educational qualification. For UKB, this filtering was not necessary since the individuals in the cohort were already older than 30 at the time of measurement (individuals born between 1930 - 1970).

- In addition to EA, we examined the relationship between bipolar disorder PGS and academic performance, under the assumption that we would also see a positive association for academic performance, especially since academic performance is typically measured before the typical age of diagnosis of bipolar disorder.
- As a sensitivity check for analysis 1, we tested the association between bipolar disorder PGS and EA with bipolar disorder diagnosis as a covariate. This analysis evaluated whether the association between bipolar disorder PGS and EA persisted after accounting for the potential impact of a bipolar disorder diagnosis on EA.

*Analysis 2*

- The pre-registration specified that academic performance in Statistics Norway (SSB) will be indexed using national test scores at age 13. This was intended to be comparable to the performance scores available in Statistics Netherlands (CBS) at age 12. This analysis in SSB was expanded to include national test scores from ages 10 and 14 in addition to age 13 to examine the longitudinal academic trajectory.
- In both SSB and CBS, we analysed individual subject test scores in addition to the total/combined scores to identify subject-specific contribution to the association between academic performance and bipolar disorder diagnosis. In particular, the individual subject scores available were Math, Reading (in Norwegian), and English in SSB, and Math, Reading, and Language (in Dutch) in CBS.
- Case Definition for Bipolar Disorder: Diagnostic information in Norway was obtained from the Norwegian Control and Payment of Health Reimbursements Database (KUHR). This register contains reimbursement data from varied sources such as primary healthcare, hospitalizations, and specialist mental health services, and so diagnosis is typically recorded multiple times from multiple sources. To increase diagnostic validity, individuals were classified as cases only if they had received a bipolar disorder diagnosis on at least five separate occasions in healthcare records. This criteria helped exclude those with transient or provisional diagnoses. This criteria was not possible in CBS since diagnosis is typically recorded only once at the start of the treatment trajectory (see Supplementary Methods 5 for more information on the healthcare systems in Norway and the Netherlands).
- The period of observation for bipolar disorder was limited to 30 years of age in SSB and 25 years of age in CBS due to the availability of diagnostic data. To account for individuals who had not received a diagnosis by the end of the observation period, we used a Cox proportional hazards model instead of the logistic regression specified in the pre-registration.
- In addition to the main model examining the association between academic performance across ages 10 – 14 and bipolar disorder diagnosis, we examined an adjusted model wherein we regressed out the previous test scores and correlated effects. This model specifies how the relative change in academic performance is related to bipolar disorder diagnosis, above and beyond an individual's academic performance at previous ages.

**Supplementary Methods 3: Data Cleaning Procedures**

*Identification of siblings:* When registry data was available (i.e., SSB and CBS) and when linkage with the genotyped cohorts were possible (i.e., with MoBa), we identified siblings as those who share the same legal mother and father. In UKB, sibling relationships were inferred using genetic relatedness metrics, namely *kinship coefficient* and *zero identity-by-state (IBS0)*.[1,2] These variables are available in UKB as a part of the genotype data. Kinship coefficient denotes the probability that two alleles, sampled at random from the same autosomal locus in two different individuals are identical by descent (IBD), or in other words, inherited from a common ancestor. For example, parent-offspring pairs have a kinship coefficient of ~0.25 since it is the probability that a random allele in the child is inherited from one specific parent (0.5) multiplied by the probability that an allele inherited by the parent from an ancestor is in turn passed to the child (0.5, independent of the first probability). Therefore, first-degree relatives, which includes parent-offspring pairs and siblings, are defined as pairs that have a pairwise kinship coefficient of ~0.25 (or more specifically, ≥ 0.177 and ≤ 0.354 to allow for normal variation). To differentiate between parent-offspring pairs and siblings, we use the zero identity-by-state (IBS0) metric, which denotes the number of genetic variants where two individuals do not share any alleles, independent of shared ancestry. Parent-offspring pairs, by definition, always share one allele in each autosomal position inherited by descent (IBD), so their IBS0 is ~ 0. On the other hand, siblings usually have a higher pairwise IBS0. We therefore define sibling pairs as those who have a zero identity-by-state (IBS0) > 0.001.[3]

*Data Filtering Process*: **Supplementary Table 1** highlights the data filtering process and the sample sizes remaining at each stage. First, duplicate records, if present, were removed, and the sample was filtered to only those who had data available on sociodemographic and parental information. For each analysis, participants were further restricted to the relevant birth cohorts and required to have complete data on the exposure, outcome, and covariates, which resulted in the final population sample. For between and within-family analyses, siblings were identified using parental identifiers or genetic relatedness metrics. Additional exclusions were applied to remove families with complex parental structures (siblings with more than one half sibling), individuals with inconsistent parental information, and those with same-sex parents. We also removed siblings who were born less than six months apart, while retaining twins and higher-order multiples born on the same day. Sibships reduced to a single individual after these filtering steps were excluded. This resulted in the final sibling sample.

**Supplementary Methods 4: UK Biobank (UKB) Genotype Data Quality Control**

Genotype data collection and imputation procedures can be found at Bycroft et al. (2018).[4] All subsequent genotype quality control procedures were carried out in the UKB Research Analysis Platform (RAP). The genotype files were first cleaned to include only the set of 1,252,123 autosomal HapMap3 (HM3) SNPs with a minor allele frequency (MAF) of > 0.01, and missingness < 0.05. We determined genetic ancestries of the sample using Principal Component Analysis (PCA) in GCTA (Yang et al., 2011). First, we extracted HM3 SNPs with MAF > 0.01 from the 1000 Genomes Phase 3 reference dataset.[5] Then we computed PCs for this reference dataset for 2,504 participants and projected the UKB individuals onto these

PCs to identify genetic ancestries. UKB individuals were assigned to different ancestries based on a >0.9 posterior probability of belonging to the 1000 Genomes reference cluster. A total of 456,566 individuals were identified to be of European ancestry. More information on ancestry classification can be found elsewhere.[6] We implemented QC procedures again in a sample of unrelated individuals of European ancestry using filters for MAF > 0.01, Hardy–Weinberg equilibrium (HWE) P value < $10^{-10}$ and missingness < 0.05. Unrelated individuals were extracted using the King software for kinship-based relationship inference using the algorithm --unrelated.[1] The set of SNPs filtered from this step were then extracted from the complete list of individuals of European ancestry. The PGS for bipolar disorder were calculated based on this cleaned genotype data.

**Supplementary Methods 5: Measures**

*I. Bipolar Disorder PGS*

In UKB, PGS were computed within the UK Biobank Research Analysis Platform (RAP). Summary statistics excluding UKB and 23andMe data is publicly available at https://figshare.com/articles/dataset/bip2024/27216117. For analysis using MoBa data, summary statistics from the most recent GWAS (O'Connell et al., 2025) excluding MoBa and 23andMe data were obtained from the first author on request.

*II. Educational Attainment (EA)*

For MoBa, detailed documentation of the Norwegian education codes (NUS) classification and its conversion to ISCED is provided by Statistics Norway: http://www.ssb.no/en/utdanning/norwegian-standard-classification-of-education. EA in UKB was derived from self-reported highest qualification (UKB Field ID: 6138). Participants reported their highest qualification achieved, with 8 possible categories including 'Prefer not to answer'. Those who preferred not to answer were excluded from analyses. If data were available from more than one instance, we used the last-known answer as their qualification.

In UKB, the highest qualification is 'University or College Degree' and offers no distinction between the type of University Degree. On the other hand, the educational qualification data available in MoBa ranged from primary education to university-level qualifications, with detailed distinctions between bachelor's, master's, and doctoral degrees, thereby providing a higher level of granularity. Additionally, UK qualifications such as NVQ, HND, or HNC represent vocational or professional training that lies between upper secondary education and a full bachelor's degree. Previous studies using UKB data have classified these qualifications as equivalent to ISCED level 6, while "college or university degree" has been mapped to ISCED level 7.[7,8] Although this classification does not perfectly align with the Norwegian system or ISCED 2011 definitions, we adopted this approach to ensure comparability with prior research (**Supplementary Table 2**).

#### *III. Academic Performance*

In SSB, the national tests taken at ages 10, 13, and 14 are compulsory, with over 96% of Norwegian students taking them. Exemptions are granted for students with special needs and those taking introductory language courses. Test results are used to monitor academic progress and have no direct consequences for students. To clean the national test scores, we removed those with 0 as their test score as this indicates non-attendance. For the students who retook the tests in different years, we used the scores of the first attempt since this is likely the test they took at the standard test age, our age of interest.

Academic performance in CBS is indexed using test results from the nationally-administered test at age 12. Up until 2015, this test was provided by CITO (https://cito.nl/) and was mandatory for all students at the end of primary school. The CITO test is typically administered at the end of elementary school. Until 2015, this nationally standardized assessment played a key role in secondary school tracking. Contrast to Norway, the results of this test, in part, determine the secondary educational track that students will pursue (e.g., academic versus technical) and therefore is of great importance. Inspection of the CITO dataset revealed that some individuals had taken the test outside the standard age range. For example, in 2006, test participants included individuals born between 1990 and 1997. Given that the CITO test is typically administered at ages 11–12, we restricted the sample to individuals aged 10–14 at the time of testing to ensure comparability. The total score provided to each student by CBS ranges from 501 to 550, calculated using a formula that adjusts for variation in test difficulty across years.

#### *IV. Bipolar Disorder Diagnosis*

***Health care systems***

Norway is a social democratic welfare state which has a universal health care system providing complete population coverage. Consequently, private health care is relatively small, with private institutions providing about 13% of all specialist mental health care and private insurance accounting for <1% of total health spending.[9] Therefore, most individuals undergoing treatment for mental health issues in Norway are captured by the registry data, making it highly reliable for examining the epidemiology of mental health outcomes.

We obtained diagnostic data from the Control and Payment of Health Reimbursements Database (KUHR), which contains all relevant information including administrative, medical and sociodemographic details. KUHR (https://helsedata.no/en/forvaltere/norwegian-directorate-of-health/norwegian-control-and-payment-of-health-reimbursements-database-kuhr/) contains bills from primary healthcare services provided by general practitioners (GPs), specialist healthcare services, and emergency outpatient clinics, among others. Primary care is generally provided by the GPs, who refer to specialists for more complex care, such as severe mental health issues.

Similar to Norway, the Netherlands is also a social welfare state, but provides universal healthcare through a heavily regulated, mandatory private insurance system. Basic health care insurance, which covers all regular and short-term medical treatments, is compulsory and available to all Dutch residence since the emergence of the new Health Insurance Act in 2006.

The Dutch mental health care system (Geestelijke Gezondheidszorg GGZ) has the first and second line of care, apart from the General Practitioner (GP), who is typically the first point of contact in the Dutch healthcare system. GPs treat simple mental health issues, which do not require a diagnosis or specialised therapy. For complex problems, the patient is referred to the first line of care, also known as basic mental health care. Basic care is provided in an outpatient setting for patients who have a single well-defined disorder. Basic care is limited to a short duration of a maximum of 8 weeks. If mental health stabilizes from basic care, further management of mental health symptoms is taken care of by general medicine.

If further mental health care is required, the patient is referred to the second line of care, also known as specialised care, intended for those with complex or severe mental health issues. When a patient enters specialised care, a "Diagnose Behandeling Combinatie (DBC)" is registered, which specifies the primary diagnosis, additional secondary diagnoses if applicable, and the treatment plan. We use the primary and secondary diagnosis recorded by these specialist services as a DBC, based on the Diagnostic and Statistical Manual of Mental Disorders 4th Edition (DSM-IV). More information about the healthcare system in the Netherlands is available at https://english.zorginstituutnederland.nl/about-us/healthcare-in-the-netherlands.

***Diagnostic Criteria***

The diagnostic information obtained from KUHR were based on the International Classification of Primary Care version 2 (ICPC-2) in primary care and International Classification of Diseases, Tenth Revision (ICD-10) in specialist care. Bipolar disorder diagnosis in KUHR was established using the ICD-10 code F31 (bipolar affective disorder) and ICPC-2 code P73.02 (Bipolar disorder / manic-depression). Diagnoses in CBS were based on the following DSM-IV codes

- 296.00 – 296.06: Bipolar I Disorder, Single Manic Episode (with specifiers for severity, partial, or full remission)
- 296.40 – 296.46: Bipolar I Disorder, Most Recent Episode Manic (with specifiers for mild, moderate, severe, or remission states)
- 296.50 – 296.56: Bipolar I Disorder, Most Recent Episode Depressed (specifiers for severity or remission states)
- 296.60 – 296.66: Bipolar I Disorder, Most Recent Episode Mixed (featuring a mix of manic and depressive symptoms simultaneously)
- 296.70: Bipolar I Disorder, Most Recent Episode Unspecified
- 296.80: Bipolar Disorder Not Otherwise Specified (NOS)
- 296.89: Bipolar II Disorder

***Age at Diagnosis***

In SSB, age at diagnosis was derived from the date of diagnosis variable. In CBS, the exact date of diagnosis was not available for the primary diagnoses, so we used date of the start of the treatment care trajectory, which was the closest possible alternative. Diagnoses may have occurred earlier or later, particularly for returning patients or those transferred from another institution.

***Period of Follow-up***

The period of follow-up for each birth cohort in SSB and CBS are presented in **Supplementary Table 3**. In SSB, diagnostic data was available between 2006 and 2025, so we followed up individuals up to a maximum of 32 years of age. For our main analyses using CBS data, we extracted diagnoses from the 'detailed' datasets GGZDBCTRAJECTENTAB and GGZDBCDIAGNOSETAB, which covers diagnoses recorded between 2011 and 2016. These datasets provide the most detailed diagnostic information, allowing precise identification of bipolar disorder cases. However, our Dutch cohort included people born between the years 1992 to 2001, restricting the maximum age at follow-up to 24 years.

Datasets with diagnostic information beyond 2016 are provided as a part of the 'hoofd' datasets (GGZDBCTRAJECTENHOOFDDIAGTAB and GGZDBCDIAGNOSEHOOFDDIAGTAB), covering diagnoses from 2015–2021. However, these datasets only include diagnoses under the broad category of "bipolar and other mood disorders," without distinguishing bipolar disorder from substance-induced mood disorders, among others. Similarly, the 'vektis' (GGZDECLVEKTIS) dataset, which provides diagnoses extracted from health insurance claims between the years 2013 and 2021, also records diagnoses at the level of "bipolar and other mood disorders" and cannot distinguish cases of bipolar disorder. An overview of all CBS datasets used in this project are provided in **Supplementary Table 4**. Sensitivity analysis using data from these other sources are outlined in **Supplementary Results 9**.

### *V. Covariates*

Key variables used in UKB were sex (UKB ID: 31), year of birth (UKB ID: 34), and birth order (derived from number of older siblings - UKB ID: 5057). The variables used in SSB included individual ID (w19_0634_ln3), sex (kjoenn), and birth date (foedsels_aar_mnd). In CBS, individual ID (RINPERSOON), sex, and year of birth (GBAGEBOORTEJAAR) were used. For models involving registry data, birth order was assigned by grouping families and ordering them based on their birth year. Multiples were assigned the same birth order. Since complete families are not recruited in UKB, birth order was derived from the variable 'number of older siblings'. This data was collected at 4 different time points and we prioritized the last available measurement.

**Supplementary Methods 6: Power Simulations for Analysis 1**

Prior to any analysis, we simulated data to compare two possible analytical methods for testing the hypothesis that bipolar disorder and EA have a positive genetic relationship. The possible options were: 1. Using EA PGS to predict bipolar disorder or alternatively, 2. Using bipolar disorder PGS to predict EA. We simulated data to compare the power of the two analyses options using the following parameters.

- Sample size (n) was set as 40,000 based on the number of sample of siblings available in the UK Biobank.
- Based on previous literature, R2 was set as 0.15 for EA PGS,[10] and 0.05 for bipolar disorder PGS.[11]
- For option 1, bipolar disorder was simulated as a dichotomous outcome with case prevalence of 1%. For option 2, EA was considered a continuous outcome.
- Because of the nature of the outcome variables, logistic and linear regressions were used for option 1 and 2 respectively.

$$Y = \beta 0 + \beta 1X + \varepsilon$$

with Y = outcome, β0 = intercept, X = predictor, and ε = error term

- The analyses were repeated 1000 times.

The linear regression of EA on bipolar disorder PGS yielded more power than the logistic regression of bipolar disorder diagnosis on EA PGS (~99% versus ~55% respectively). As a result, we decided to use bipolar disorder PGS as the predictor and EA continuous variable as the outcome in our final analyses. Code used for the simulation can be found on GitHub. https://github.com/VaishnaviMadhavan/BipolarDisorder-EA/tree/main/Simulations.

**Supplementary Results 7: Analysis 1 Educational Attainment (EA) Sensitivity Check**

As a sensitivity check, we examined whether the association between bipolar disorder PGS and educational attainment (EA) persisted after accounting for the potential impact of a bipolar disorder diagnosis on educational attainment.

In SSB, we used the diagnostic data extracted from national registers, as used for our analysis 2 and described in **Supplementary Methods 5**. In UKB, we used combination of self-report and physician-diagnosed records to ascertain bipolar disorder cases. The variables used in UKB include

- Self-reported illness: psychological/psychiatric problem: mania/bipolar disorder/manic depression (UKB ID: 20002)

- Bipolar disorder status (UKB ID: 20122)
- Bipolar and major depression status (UKB ID: 20126)
- Mental health problems ever diagnosed by a professional: Mania, hypomania, bipolar or manic-depression (UKB ID: 20544)
- Diagnoses - main ICD10: Chapter V Mental and behavioral disorders: F31 Bipolar affective disorder (F31.0-F31.9) (UKB ID: 41202)
- Diagnoses - secondary ICD10: Chapter V Mental and behavioral disorders: F31 Bipolar affective disorder (F31.0-F31.9) (UKB ID: 41204)

If an individual reported bipolar disorder in any one of the above variables, we considered them as a bipolar disorder case. We included both bipolar Type 1 (Mania) and 2 (Hypomania) disorder as well as mania and manic depression. As shown in **Supplementary Table 7**, adding bipolar disorder diagnosis as a covariate did not significantly change our results; the association between bipolar disorder PGS and EA remains unchanged in regards to the direction of association (positive), significance, and magnitude after accounting for bipolar disorder diagnostic status.

**Supplementary Results 8: Examining Non-linearity**

Previous literature have reported non-linear associations between academic performance and bipolar disorder diagnosis,[12,13] showing a "reverse J-shaped" association with a modestly elevated risk of bipolar disorder among those with academic performance > 2 SD above the mean and the highest risk of bipolar disorder among individuals with the lowest academic performance. Here, we investigate the possibility of non-linear associations between academic performance and bipolar disorder diagnosis in our datasets, SSB and CBS.

First, to visually assess the functional form of the association between academic performance and bipolar disorder diagnosis, standardized school test scores were divided into equal-width bins of 0.25 standard deviations (SD). Within each bin, the observed percentage of individuals diagnosed with bipolar disorder was calculated. These observed percentages were plotted within each bin, and a locally smoothed curve was overlaid to aid visualization of the overall trend. This graph is presented in **Supplementary Figure 3**. Based on visual inspection, it seems like English scores at age 10 as well as Reading scores at ages 13 and 14 might have a J-shaped non-linear association between academic performance and bipolar disorder diagnosis. However, it is difficult to say with certainty due to the low number of observations at the extreme ends of the Z-score distribution.

Next, as a formal test of non-linearity, we fitted Cox proportional hazards regression models in which test performance was modelled both as a continuous linear predictor and using a natural cubic spline with four degrees of freedom. The linear and spline models were fitted to the same analytical sample and included the same covariates. We compared the models using likelihood-ratio tests, with the null hypothesis that the linear model provided an adequate description of the association. A p-value below 0.05 was taken to indicate evidence of non-linearity (**Supplementary Table 11**). For visualization, we plotted the estimated hazard ratio for bipolar disorder diagnosis across the standardized test-score distribution from the spline models, with the hazard ratio at the mean test score (0 SD) used as the reference and 95% confidence intervals (**Supplementary Figure 4**).

The spline analyses provided evidence of non-linearity only for mathematics and English scores at age 10. Specifically, the likelihood-ratio test comparing the linear and spline models indicated significant non-linearity for mathematics ($\chi^2 = 8.746$, $p = 0.033$) and English ($\chi^2 = 12.171$, $p = 0.006$), out of which only English scores might have a possible "J-shaped" association. Overall, our statistical tests suggest very weak evidence of non-linear associations in our data.

**Supplementary Results 9: Case Ascertainment in Statistics Netherlands (CBS) and Follow-up Analyses**

For our main analyses, we used the 'detailed' datasets with complete diagnostic information spanning from 2011 to 2016 (**Supplementary Method 5**). As a sensitivity analysis, we checked the utility of including additional years of diagnostic information but broader diagnostic labels ('bipolar and other mood disorders') available until 2021 from the hoofd and vektis datasets. **Supplementary Table 4** provides an overview of the datasets available and their limitations. The analysis which includes diagnoses collated across all datasets is labelled 'All Datasets' in **Supplementary Figure 5**.

In addition, because of the overlap between the detailed, hoofd, and vektis datasets for the years 2015 and 2016 and the availability of prescription data, we were able to employ prediction modelling to identify which individuals identified as having 'bipolar and other mood disorders' in the hoofd and vektis datasets are likely to have bipolar disorder. Prescription records were obtained from the dataset 'MEDICIJNTAB' and combined with the high quality data from 2011-2016 to predict likely bipolar disorder cases in the latter years. Prescriptions were classified using the ATC (Anatomical Therapeutic Chemical) system, with the list of all medication classes presented in **Supplementary Table 12**.

For the development and validation of the prediction model, a three-stage framework was used to generate predicted outcome probabilities for those who only had a diagnosis of 'bipolar and other mood disorders'. The three steps included training, validation, and prediction.

*Step 1 – Training:* We first selected individuals who have a diagnosis of 'bipolar and other mood disorder' in either hoofd or vektis dataset between 2015 and 2016. This dataset was split into training and validation datasets. 80% of these individuals were randomly used to train the prediction model and the rest 20% were used for validation. The true cases of bipolar disorder in the training dataset was obtained from the detailed dataset. Then we fitted logistic regression models to use the prescription ATC classes to predict a bipolar disorder diagnosis in the detailed dataset. From this model, we extracted model predictions and their associated standard errors using the R package 'Predict'.[14] The ROC-AUC (Receiver Operating Characteristic - Area Under the Curve) comparing the predicted probabilities and the observed outcomes in the training dataset was 0.752.

*Step 2 – Validation:* The estimated regression coefficients (**Supplementary Table 12**) from the training model were then applied to the validation dataset to generate predicted probability values for each individual. This metric ranges from 0 to 1, and indicates how likely an individual has a diagnosis of bipolar disorder, with values closer to 1 indicating a high probability of being a case. Because observed outcome data were available in the validation dataset, predictive performance was assessed by comparing the predicted values with the observed outcome values. The ROC-AUC comparing the predicted probabilities and the observed outcomes in the validation dataset was 0.749. In **Supplementary Table 13**, we present the false positive and false negatives identified in the validation dataset using different cut-offs for predicted probabilities.

*Step 3 - Prediction:* The fitted model was then applied to the prediction dataset which contains prescription data but no observed bipolar disorder diagnosis between the years 2016 and 2021. Similar to the validation step, predicted probabilities were computed using the regression coefficients estimated in the training dataset. These predicted probabilities were used in two different ways: Firstly, they were added as weights in the cox proportional hazards regression model using the argument 'weights'. Secondly, we used 4 different cut-offs (0.6, 0.7, 0.8, 0.9) in the predicted probabilities to ascertain cases; individuals with predicted probabilities value below the cutoff will be considered as controls.

**Supplementary Figure 5** shows the relationship between academic performance and bipolar disorder diagnosis using the different ways of ascertaining cases. The addition of additional years of diagnostic information seems to produce results in a similar direction to the main analysis, indicating a positive association between academic performance and bipolar disorder diagnosis. The population estimates are positive and significant but the within-sibship estimates are predominantly not significant. The case ascertainment in the main model is accurate but the period of follow-up is restricted to a maximum of 24 years of age, and so the proportion of cases are low. On the other hand, inclusion of all datasets extended the period of follow-up to 30 years, but involves imprecise case ascertainment, potentially inducing measurement error. Across all the follow-up analyses, the population estimates were positive and significant, but the within-sibship estimate was mostly not significant. The estimates in our follow-up analyses have smaller confidence intervals at the 95% level compared to the main analysis, but the attenuation could possibly indicate measurement error due to the inclusion of individuals with diagnoses other than bipolar disorder.

**Supplementary Figure 1. Associations between bipolar disorder PGS and academic performance at ages 10, 13, 14.**

*MoBa = The Norwegian Mother, Father, and Child Cohort Study; SSB = Statistics Norway. Reading comprehension test was administered in Norwegian. Pink represents the population estimate, green and blue represent the between-sibship and within-sibship estimates respectively obtained from the sibling sample. The lines represent 95% confidence intervals.*

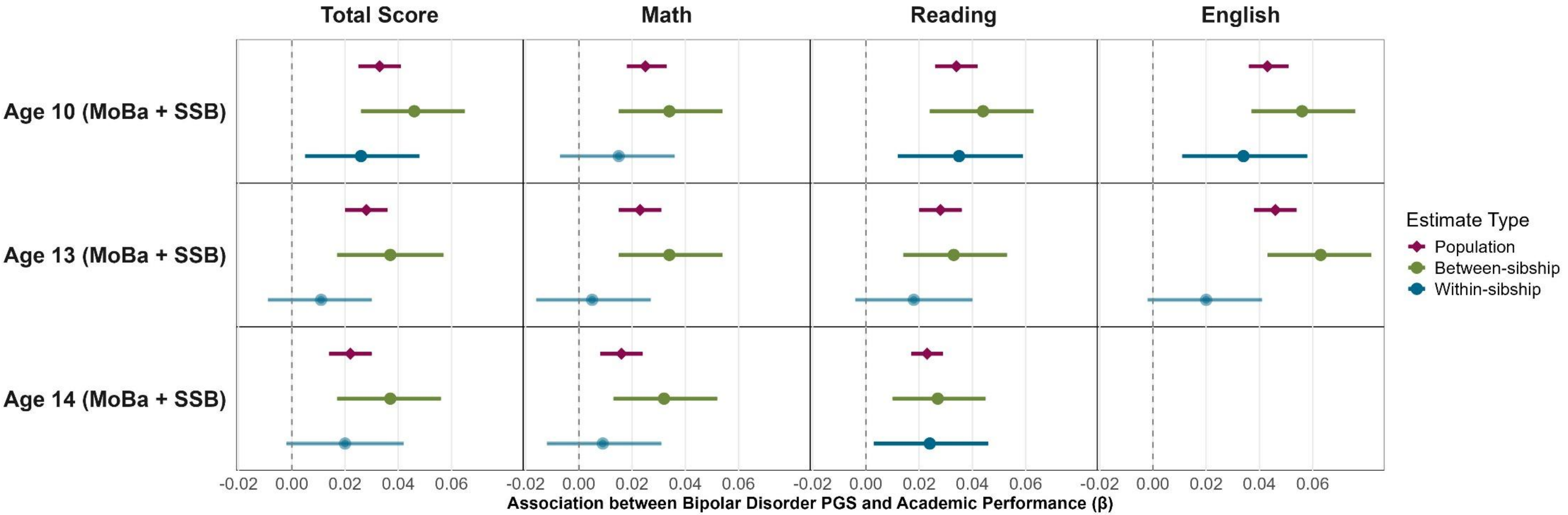

**Supplementary Figure 2. Associations between academic performance and bipolar disorder diagnosis.**

*The estimates at ages 10, 13, and 14 are from Statistics Norway (SSB) and the estimates at age 12 are from Statistics Netherlands (CBS). Reading comprehension tests are administered in Norwegian in SSB and Dutch in SSB. Pink represents the population estimate, green and blue represent the between-sibship and within-sibship estimates respectively obtained from the sibling sample. The lines represent 95% confidence intervals; these are not necessarily symmetric around the estimate as hazard ratios from Cox regression are estimated on the log scale and then exponentiated.*

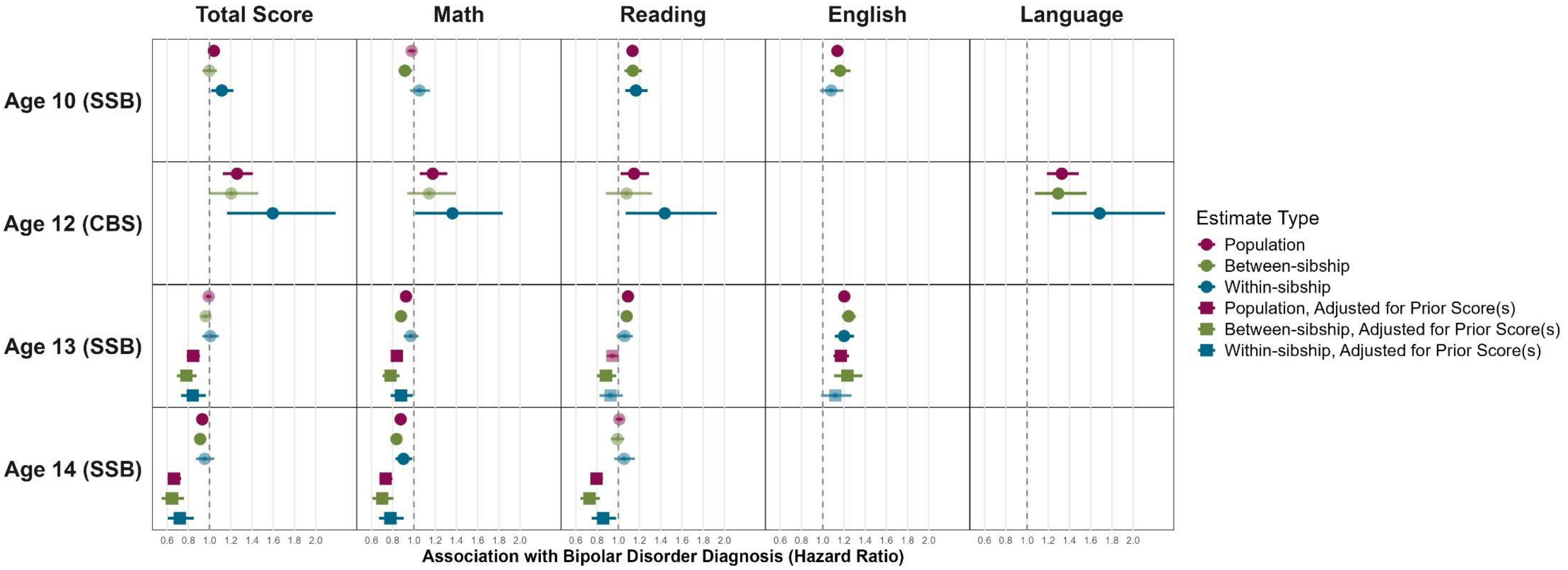

**Supplementary Figure 3. Observed percentage of bipolar disorder diagnoses across standardized school test scores.**

*Scores from ages 10, 13, and 14 were obtained from Statistics Norway (SSB), whereas the age 12 scores were obtained from Statistics Netherlands (CBS). School test scores were divided into 0.25 standard deviation (SD) intervals, and the observed percentage of individuals diagnosed with bipolar disorder was calculated within each interval. The blue curve represents a locally smoothed trend to facilitate visual assessment of the functional form of the association. Panels show figures separately by test and age.*

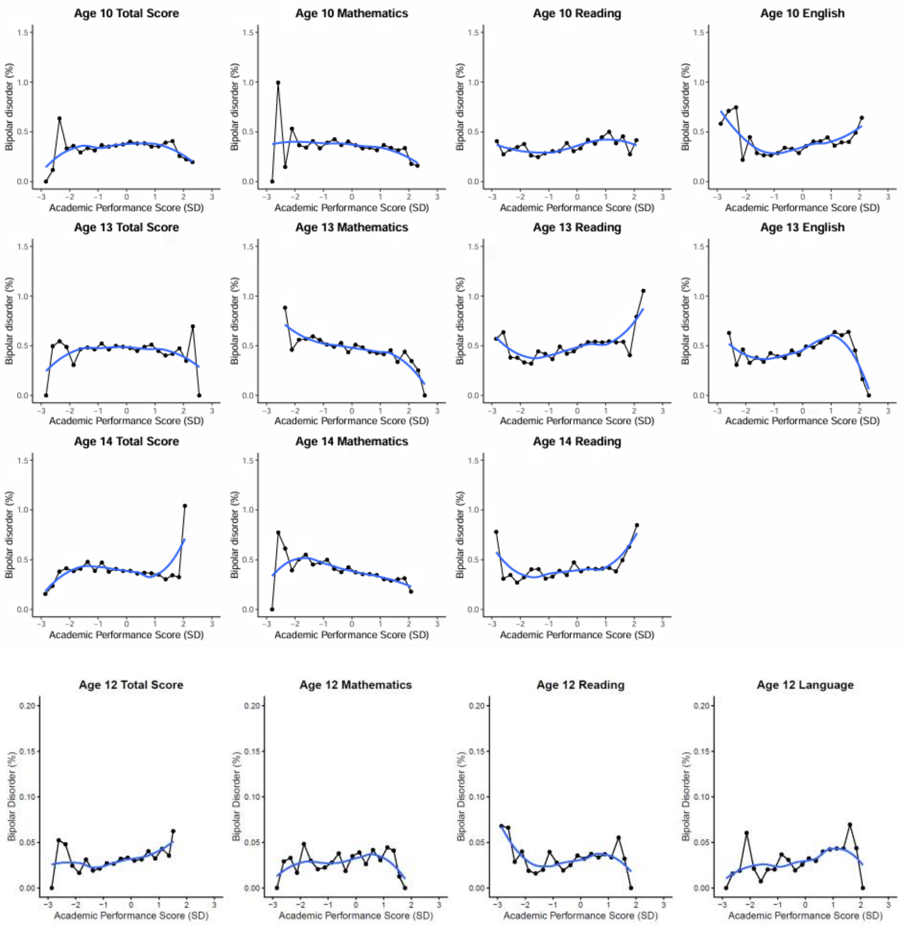

**Supplementary Figure 4. Non-linear associations between school test performance and bipolar disorder diagnosis.**

*Curves show hazard ratios (HR) for bipolar disorder diagnosis across the distribution of standardized school test scores, estimated using Cox proportional hazards models with natural cubic splines. HRs are shown relative to the mean test score (0 SD). Shaded areas represent 95% confidence intervals. The dashed horizontal line indicates a hazard ratio of 1. The vertical dotted line indicates the mean standardized test score. Panels show associations separately by test and age.*

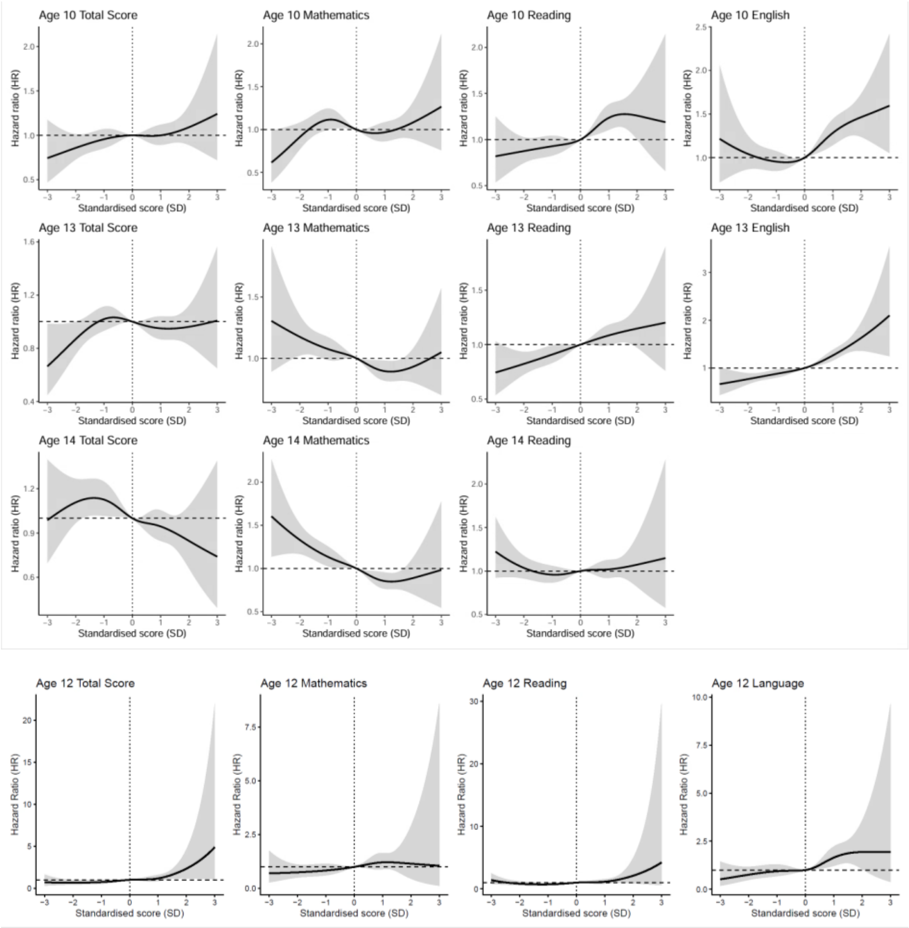

**Supplementary Figure 5. Comparison of different methods of case ascertainment in Statistics Netherlands (CBS).**

*'Main Model' uses diagnostic information from detailed dataset covering diagnoses from 2011 to 2016. 'All Datasets' uses diagnostic information from detailed, hoofd and vektis datasets, spanning from 2011 to 2021, with imprecise case ascertainment from 2017. 'All Datasets Weighted' uses predicted probabilities as weights in the cox proportional hazards regression model. The 'Cutoff' estimates use the predictive probabilities to ascertain cases using a threshold. Pink represents the population estimate, green and blue represent the between-sibship and within-sibship estimates respectively obtained from the sibling sample. The lines represent 95% confidence intervals.*

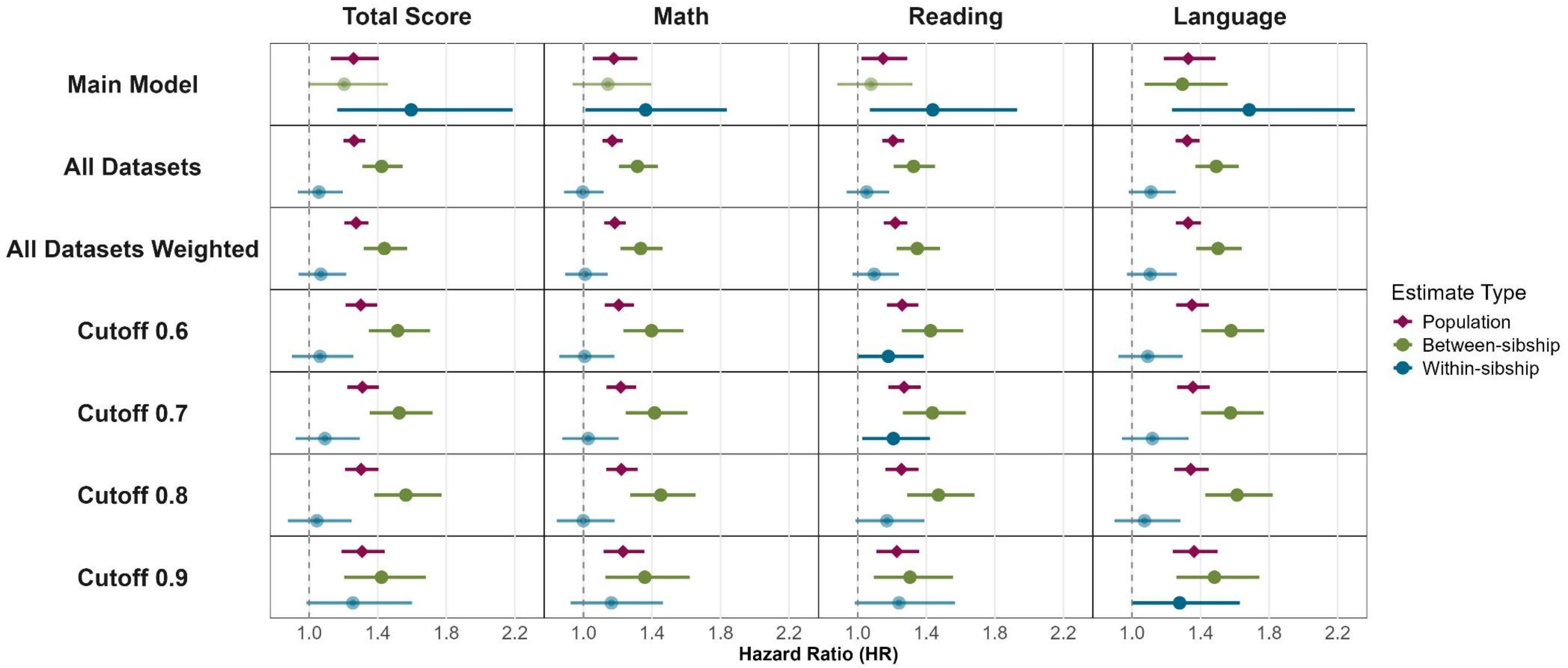

**Supplementary Table 1. Data filtering, separated by analysis and cohort.**

*MoBa = the Norwegian Mother, Father and Child Cohort Study; UKB = UK Biobank; SSB = Statistics Norway; CBS = Statistics Netherlands*

| Sample Filtering | Analysis 1 | | Analysis 2 | |
|---|---|---|---|---|
| | SSB + MoBa (Norway) | UKB (UK) | SSB (Norway) | CBS (Netherlands) |
| **Population Data** | | | | |
| Original Dataset | 9,080,159 | 192,726 | 9,080,159 | 6,726,695 |
| Removed Duplicated Records | 9,080,090<br>(n=69 removed) | 192,726 | 9,080,090<br>(n=69 removed) | 6,726,695 |
| Removed missing sociodemographic/parental information | 5,038,126<br>(n=4,041,964 removed) | 192,726 | 5,038,126<br>(n=4,041,964 removed) | 4,256,591<br>(n=2,470,104 removed) |
| Total Population Sample with Data Available on All Relevant Variables | **121,550** | **192,726** | **1,026,024** | **1,144,287** |
| **Sibling Data** | | | | |
| Population data with valid sociodemographic/parental information | 5,038,126 | 192,726 | 5,038,126 | 4,256,591 |
| Siblings with valid parental information | 4,269,661<br>(n=768,465 removed) | 192,726 | 4,269,661<br>(n=768,465 removed) | 3,324,523<br>(n=932,068 removed) |
| Removed individuals whose parents had more than one child with another partner | 3,101,512<br>(n=1,168,149 removed) | 192,726 | 3,101,512<br>(n=1,168,149 removed) | 3,274,878<br>(n=49,645 removed) |
| Removed those with same-sex parents | 3,099,830<br>(n=1,682 removed) | 192,726 | 3,099,830<br>(n=1,682 removed) | 3,271,554<br>(n=3,324 removed) |
| Removed parents with mismatching information (birthday and land of origin) | 3,099,830 | 192,726 | 3,099,830 | 3,266,658<br>(n=5,465 removed) |

| | | | | |
|---|---|---|---|---|
| Removed siblings born less than 6 months apart (but not twins and multiples) | 3,099,055 (n=775 removed) | 192,726 | 3,099,055 (n=775 removed) | 3,266,089 (n=569 removed) |
| Sibling Sample with Data Available on All Relevant Variables | 16,746 nested within 8,107 families | 12,284 nested within 5,997 families | 496,969 nested within 208,350 families | 619,796 nested within 286,658 families |

**Supplementary Table 2. Conversion of educational codes into ISCED 2011 and years of education (EA).**

*MoBa = the Norwegian Mother, Father and Child Cohort Study; UKB = UK Biobank; NUS = Norwegian education codes; ISCED = International Standard Classification of Education*

| MoBa | | UKB | | ISCED 2011 | Years of Education (EA) |
|---|---|---|---|---|---|
| **NUS 2000** | **NUS 2000 Label** | **UKB Code** | **UKB Label** | | |
| 0 | No Education | | | 0 | 0 |
| 1 | Primary Education | -7 | None of the Above | 1 | 7 |
| 2 | Lower Secondary Education | 3 | O levels/GCSEs or equivalent | 2 | 10 |
| | | 4 | CSEs or equivalent | | |
| 3 | Upper Secondary Education, Basic | 2 | A levels/AS levels or equivalent | 3 | 13 |
| 4 | Upper Secondary Education, Final Year | | | | |
| 5 | Post-Secondary Non-tertiary Education | 6 | Other prof. qual. e.g.: nursing, teaching | 5 | 15 |
| 6 | Undergraduate Education (Bachelor's) | 5 | NVQ or HND or HNC or equivalent | 6 | 19 |
| 7 | Graduate Education (Master's) | 1 | College or University degree | 7 | 20 |
| 8 | Postgraduate Education (PhD) | - | - | 8 | 22 |

**Supplementary Table 3. Period of Education and Diagnostic Data Available in Statistics Norway (SSB) and Statistics Netherlands (CBS).**

*The table displays the birth cohorts for SSB and CBS, years of education and diagnostic data available, the age at which academic performance was measured (pink boxes), and the period of diagnostic follow-up for each birth cohort (green boxes). In CBS, academic performance is typically measured at age 12, however, children aged 10–14 were included, due to variation in test-taking age across birth cohorts. There was similar variation at each age in SSB.*

| **Statistics Norway (SSB)** | | | | | | | | | | | | | | | | | | | | |
|---|---|---|---|---|---|---|---|---|---|---|---|---|---|---|---|---|---|---|---|---|
| Years of Education Data Available | | 2007 | 2008 | 2009 | 2010 | 2011 | 2012 | 2013 | 2014 | 2015 | 2016 | 2017 | 2018 | 2019 | 2020 | 2021 | 2022 | 2023 | 2024 | 2025 |
| Years of Diagnosis Data Available | 2006 | 2007 | 2008 | 2009 | 2010 | 2011 | 2012 | 2013 | 2014 | 2015 | 2016 | 2017 | 2018 | 2019 | 2020 | 2021 | 2022 | 2023 | 2024 | 2025 |
| Birth Year | Age | | | | | | | | | | | | | | | | | | | |
| 1993 | 13 | 14 | 15 | 16 | 17 | 18 | 19 | 20 | 21 | 22 | 23 | 24 | 25 | 26 | 27 | 28 | 29 | 30 | 31 | 32 |
| 1994 | 12 | 13 | 14 | 15 | 16 | 17 | 18 | 19 | 20 | 21 | 22 | 23 | 24 | 25 | 26 | 27 | 28 | 29 | 30 | 31 |
| 1995 | 11 | 12 | 13 | 14 | 15 | 16 | 17 | 18 | 19 | 20 | 21 | 22 | 23 | 24 | 25 | 26 | 27 | 28 | 29 | 30 |
| 1996 | 10 | 11 | 12 | 13 | 14 | 15 | 16 | 17 | 18 | 19 | 20 | 21 | 22 | 23 | 24 | 25 | 26 | 27 | 28 | 29 |
| 1997 | 9 | 10 | 11 | 12 | 13 | 14 | 15 | 16 | 17 | 18 | 19 | 20 | 21 | 22 | 23 | 24 | 25 | 26 | 27 | 28 |
| 1998 | 8 | 9 | 10 | 11 | 12 | 13 | 14 | 15 | 16 | 17 | 18 | 19 | 20 | 21 | 22 | 23 | 24 | 25 | 26 | 27 |
| 1999 | 7 | 8 | 9 | 10 | 11 | 12 | 13 | 14 | 15 | 16 | 17 | 18 | 19 | 20 | 21 | 22 | 23 | 24 | 25 | 26 |
| 2000 | 6 | 7 | 8 | 9 | 10 | 11 | 12 | 13 | 14 | 15 | 16 | 17 | 18 | 19 | 20 | 21 | 22 | 23 | 24 | 25 |
| 2001 | 5 | 6 | 7 | 8 | 9 | 10 | 11 | 12 | 13 | 14 | 15 | 16 | 17 | 18 | 19 | 20 | 21 | 22 | 23 | 24 |
| 2002 | 4 | 5 | 6 | 7 | 8 | 9 | 10 | 11 | 12 | 13 | 14 | 15 | 16 | 17 | 18 | 19 | 20 | 21 | 22 | 23 |
| 2003 | 3 | 4 | 5 | 6 | 7 | 8 | 9 | 10 | 11 | 12 | 13 | 14 | 15 | 16 | 17 | 18 | 19 | 20 | 21 | 22 |
| 2004 | 2 | 3 | 4 | 5 | 6 | 7 | 8 | 9 | 10 | 11 | 12 | 13 | 14 | 15 | 16 | 17 | 18 | 19 | 20 | 21 |
| 2005 | 1 | 2 | 3 | 4 | 5 | 6 | 7 | 8 | 9 | 10 | 11 | 12 | 13 | 14 | 15 | 16 | 17 | 18 | 19 | 20 |
| 2006 | | 1 | 2 | 3 | 4 | 5 | 6 | 7 | 8 | 9 | 10 | 11 | 12 | 13 | 14 | 15 | 16 | 17 | 18 | 19 |
| 2007 | | | 1 | 2 | 3 | 4 | 5 | 6 | 7 | 8 | 9 | 10 | 11 | 12 | 13 | 14 | 15 | 16 | 17 | 18 |
| 2008 | | | | 1 | 2 | 3 | 4 | 5 | 6 | 7 | 8 | 9 | 10 | 11 | 12 | 13 | 14 | 15 | 16 | 17 |
| 2009 | | | | | 1 | 2 | 3 | 4 | 5 | 6 | 7 | 8 | 9 | 10 | 11 | 12 | 13 | 14 | 15 | 16 |
| 2010 | | | | | | 1 | 2 | 3 | 4 | 5 | 6 | 7 | 8 | 9 | 10 | 11 | 12 | 13 | 14 | 15 |

| Statistics Netherlands (CBS) | | | | | | | | | | | |
|---|---|---|---|---|---|---|---|---|---|---|---|
| Years of Education Data Available | 2006 | 2007 | 2008 | 2009 | 2010 | 2011 | 2012 | 2013 | 2014 | | |
| Years of Diagnosis Data Available | | | | | | 2011 | 2012 | 2013 | 2014 | 2015 | 2016 |
| Birth Year | Age | | | | | | | | | | |
| 1992 | 14 | 15 | 16 | 17 | 18 | 19 | 20 | 21 | 22 | 23 | 24 |
| 1993 | 13 | 14 | 15 | 16 | 17 | 18 | 19 | 20 | 21 | 22 | 23 |
| 1994 | 12 | 13 | 14 | 15 | 16 | 17 | 18 | 19 | 20 | 21 | 22 |
| 1995 | 11 | 12 | 13 | 14 | 15 | 16 | 17 | 18 | 19 | 20 | 21 |
| 1996 | 10 | 11 | 12 | 13 | 14 | 15 | 16 | 17 | 18 | 19 | 20 |
| 1997 | 9 | 10 | 11 | 12 | 13 | 14 | 15 | 16 | 17 | 18 | 19 |
| 1998 | 8 | 9 | 10 | 11 | 12 | 13 | 14 | 15 | 16 | 17 | 18 |
| 1999 | 7 | 8 | 9 | 10 | 11 | 12 | 13 | 14 | 15 | 16 | 17 |
| 2000 | 6 | 7 | 8 | 9 | 10 | 11 | 12 | 13 | 14 | 15 | 16 |
| 2001 | 5 | 6 | 7 | 8 | 9 | 10 | 11 | 12 | 13 | 14 | 15 |

**Supplementary Table 4. Overview of datasets used from Statistics Netherlands (CBS).**

| Type of Data | Name of the Dataset | Notes | Years of Available Data |
|---|---|---|---|
| Municipal registration with birth details | GBAPERSOONTAB | This dataset contains demographic and birth-related information for all registered individuals in the Netherlands. Each individual is assigned a unique identifier, ‘RINPERSOON’, which enables linkage across datasets. | .. |
| Legal parent-child linkage data | KINDOUDERTAB | This dataset contain information on legal parent-child relationships, allowing identification of family structures. We used this dataset to identify siblings based on those sharing the same legal parents. | |

| | | | |
|---|---|---|---|
| Academic Performance Scores | CITOTAB | This dataset provides information on academic performance scores from the CITO test, taken at approximately age 12. We obtained scores on individual tests and aggregate scores. Data are available for the period 2006 to 2021. | 2006 - 2014 |
| Diagnostic Data | | | |
| Primary Diagnoses from Mental Health Care Trajectories | GGZDBCTRAJECTENTAB | This include treatment care trajectories of individuals utilising mental health care between 2011 and 2016. Prior to 2014, this concerned secondary mental health care, and from 2014 onwards, it includes treatments with specialised mental health care. Several diagnosis based on DSM-IV is available, from which we extracted bipolar disorder diagnosis. | 2011 - 2016 |
| Secondary Diagnoses from Mental Health Care Trajectories | GGZDBCDIAGNOSETAB | | 2011 - 2016 |
| Primary Diagnoses from Mental Health Care Trajectories | GGZDBCTRAJECTENHOOFDDIAGTAB | Includes diagnoses categorized as “bipolar and other mood disorders.” | 2015 - 2021 |
| Secondary Diagnoses from Mental Health Care Trajectories | GGZDBCDIAGNOSEHOOFDDIAGTAB | | 2015 - 2021 |
| Health Insurance Declarations | GGZDECLVEKTIS | All forms of mental health care reimbursed under the Health Insurance Act and under supplementary insurance. Data are recorded under the category ‘Bipolar and other mood disorders’. | 2013 - 2021 |
| Prescription Data | MEDICIJNTAB | Dispensed medicines are classified into ATC classes (where ATC stands for Anatomic, Therapeutic, Chemical), according to the WHO (World Health Organization) drug classification system. | 2006 - 2024 |

**Supplementary Table 5. Full descriptive table.**

MoBa = the Norwegian Mother, Father and Child Cohort Study; SSB = Statistics Norway; CBS = Statistics Netherlands. The descriptives are presented for the total sample and segregated by data available at ages 10, 13, and 14. The sibling sample includes families with at least 2 siblings.

| Analysis 1: Bipolar Disorder PGS and Educational Outcomes (EA and Academic Performance) | | | | | | | | |
|---|---|---|---|---|---|---|---|---|
| | MoBa + SSB | | | | | | | |
| | Total Sample | | Age 10 | | Age 13 | | Age 14 | |
| Variable | Population Sample | Sibling Sample | Population Sample | Sibling Sample | Population Sample | Sibling Sample | Population Sample | Sibling Sample |
| Sample Size | 74,104 | 19,091 | 72,923 | 18,616 | 73,136 | 18,689 | 71,956 | 18,845 |
| Number of Families | 64,290 | 9,277 | 63,362 | 9,055 | 63,532 | 9,085 | 62,613 | 9,158 |
| Birth Year Range | 1999-2009 | 1999-2009 | 1999-2009 | 1999-2009 | 1999-2009 | 1999-2009 | 1999-2009 | 1999-2009 |
| Females, n(%) | 36368 (49.08%) | 9354 (49%) | 35948 (49.3%) | 9175 (49.29%) | 35968 (49.18%) | 9181 (49.13%) | 35381 (49.17%) | 9247 (49.07%) |
| Bipolar Disorder PGS, M(SD) | 2.76 (0.6) | 2.76 (0.6) | 2.76 (0.6) | 2.75 (0.6) | 2.76 (0.6) | 2.75 (0.6) | 2.76 (0.6) | 2.75 (0.6) |
| Age 10 Total Score, M(SD) | 22.07 (7.2) | 22.42 (7.2) | 22.07 (7.2) | 22.42 (7.2) | - | - | - | - |
| Age 10 Math, M(SD) | 26.31 (9.1) | 27.18 (9) | 26.31 (9.1) | 27.22 (8.97) | - | - | - | - |
| Age 10 Reading, M(SD) | 20.53 (6.31) | 20.86 (6.32) | 20.53 (6.31) | 20.89 (6.31) | - | - | - | - |
| Age 10 English, M(SD) | 27.08 (10.05) | 27.14 (9.96) | 27.08 (10.05) | 27.16 (9.95) | - | - | - | - |
| Age 13 Total Score, M(SD) | 27.07 (9.53) | 27.27 (9.54) | | | 27.07 (9.53) | 27.47 (9.54) | - | - |
| Age 13 Math, M(SD) | 28.12 (10.2) | 29.22 (10.09) | - | - | 28.12 (10.2) | 29.23 (10.08) | - | - |
| Age 13 Reading, M(SD) | 27.35 (8.55) | 27.95 (8.45) | - | - | 27.35 (8.55) | 27.97 (8.44) | - | - |

| | | | | | | | | | | |
|---|---|---|---|---|---|---|---|---|---|---|
| Age 13 English, M(SD) | 28.67 (11.43) | 28.84 (11.42) | - | - | 28.67 (11.43) | 28.83 (11.41) | - | - | | |
| Age 14 Total Score, M(SD) | 29.47 (9.16) | 29.97 (9.19) | - | - | - | - | 29.47 (9.16) | 29.98 (9.18) | | |
| Age 14 Math, M(SD) | 31.02 (9.99) | 32.18 (9.87) | - | - | - | - | 31.02 (9.99) | 32.21 (9.85) | | |
| Age 14 Reading, M(SD) | 30.52 (8.26) | 31.04 (8.08) | - | - | - | - | 30.52 (8.26) | 31.07 (8.06) | | |
| Bipolar Disorder Cases | 153 (0.21%) | 33 (0.17%) | 149 (0.2%) | 32 (0.17%) | 147 (0.2%) | 30 (0.16%) | 140 (0.19%) | 31 (0.16%) | | |
| Age at Diagnosis, M(SD) | 18.09 (3.21) | 17.11 (3.73) | 18.17 (3.16) | 17.35 (3.51) | 18.26 (3.16) | 17.25 (3.79) | 18.34 (3.06) | 17.24 (3.73) | | |
| **Analysis 2: Academic Performance and Bipolar Disorder Diagnosis** | | | | | | | | | | |
| | **SSB** | | | | | | | | **CBS** | |
| | **Total Sample** | | **Age 10** | | **Age 13** | | **Age 14** | | **Age 12** | |
| **Variable** | **Population Sample** | **Sibling Sample** | **Population Sample** | **Sibling Sample** | **Population Sample** | **Sibling Sample** | **Population Sample** | **Sibling Sample** | **Population Sample** | **Sibling Sample** |
| Sample Size | 1026024 | 496969 | 807240 | 369016 | 1001462 | 483156 | 874986 | 487698 | 1,130,598 | 619,796 |
| Number of Families | 626043 | 208350 | 515908 | 159001 | 615081 | 203333 | 551183 | 204933 | 797,460 | 286,658 |
| Birth Year Range | 1993-2010 | 1993-2010 | 1994-2010 | 1994-2010 | 1993-2010 | 1993-2010 | 1994-2010 | 1993-2010 | 1992 - 2001 | 1992 - 2001 |
| Females, n(%) | 502113 (48.94%) | 242262 (48.75%) | 397627 (49.26%) | 180749 (48.98%) | 491130 (49.04%) | 235911 (48.83%) | 429366 (49.07%) | 238086 (48.82%) | 572070 (49.99%) | 309227 (49.89%) |
| Age 10 Total Score, M(SD) | 22.07 (7.2) | 22.42 (7.2) | 22.07 (7.2) | 22.42 (7.2) | - | - | - | - | - | - |
| Age 10 Math, M(SD) | 25.14 (9.63) | 25.59 (9.56) | 25.14 (9.63) | 25.59 (9.56) | - | - | - | - | - | - |
| Age 10 Reading, M(SD) | 18.83 (6.47) | 19.12 (6.5) | 18.83 (6.47) | 19.12 (6.5) | - | - | - | - | - | - |
| Age 10 English, M(SD) | 25.26 (9.8) | 25.45 (9.8) | 25.26 (9.8) | 25.45 (9.8) | - | - | - | - | - | - |

| | | | | | | | | | | |
|---|---|---|---|---|---|---|---|---|---|---|
| Age 13 Total Score, M(SD) | 27.07 (9.53) | 27.47 (9.54) | - | - | 27.07 (9.53) | 27.47 (9.54) | - | - | - | - |
| Age 13 Math, M(SD) | 27.88 (12.15) | 28.46 (12.06) | - | - | 27.88 (12.15) | 28.46 (12.06) | - | - | - | - |
| Age 13 Reading, M(SD) | 26.1 (8.94) | 26.33 (8.97) | - | - | 26.1 (8.94) | 26.33 (8.97) | - | - | - | - |
| Age 13 English, M(SD) | 26.92 (11.41) | 27.16 (11.54) | - | - | 26.92 (11.41) | 27.16 (11.54) | - | - | - | - |
| Age 14 Total Score, M(SD) | 29.47 (9.16) | 29.97 (9.19) | - | - | - | - | 29.47 (9.16) | 29.98 (9.18) | | |
| Age 14 Math, M(SD) | 29.63 (11.49) | 30.41 (11.45) | - | - | - | - | 29.63 (11.49) | 30.46 (11.43) | - | - |
| Age 14 Reading, M(SD) | 29.07 (8.91) | 29.32 (8.89) | - | - | - | - | 29.07 (8.91) | 29.35 (8.87) | - | - |
| Age 12 Total Score, M(SD) | - | - | - | - | - | - | - | - | 535.15 (9.76) | 535.94 (9.49) |
| Age 12 Math, M(SD) | - | - | - | - | - | - | - | - | 42.34 (11.14) | 43.28 (10.83) |
| Age 12 Reading, M(SD) | - | - | - | - | - | - | - | - | 29.83 (6.19) | 30.3 (6.03) |
| Age 12 Language, M(SD) | - | - | - | - | - | - | - | - | 73.63 (13.19) | 74.62 (12.91) |
| Bipolar Disorder Cases | 4995 (0.49%) | 2654 (0.53%) | 2951 (0.37%) | 1421 (0.39%) | 4898 (0.49%) | 2588 (0.54%) | 3498 (0.4%) | 2604 (0.53%) | 378 (0.03%) | 173 (0.03%) |
| Age at Diagnosis, M(SD) | 21.3 (3.83) | 21.21 (3.71) | 20.26 (3.39) | 20.11 (3.28) | 21.37 (3.78) | 21.27 (3.69) | 20.74 (3.46) | 21.24 (3.7) | 18.7 (2.56) | 18.59 (2.49) |

**Supplementary Table 6. Full regression results for analysis 1 examining the association between bipolar disorder PGS and educational attainment (EA).**

MoBa = the Norwegian Mother, Father and Child Cohort Study; SSB = Statistics Norway; UKB = UK Biobank; EA = Educational Attainment; SE = Standard Error; CI = Confidence Interval.
*p < .05. **p < .01. ***p < .001.

| Cohort | Model | Variable | Estimate | SE | 95% CI | p | Significance | N | Analysis |
|---|---|---|---|---|---|---|---|---|---|
| MoBa + SSB | Population | Intercept | 25.11 | 1.051 | [23.05, 27.17] | <0.001 | *** | 121,550 | Main Model |
| MoBa + SSB | Population | Bipolar PGS | 0.026 | 0.003 | [0.021, 0.032] | <0.001 | *** | 121,550 | Main Model |
| MoBa + SSB | Population | Sex | 0.287 | 0.005 | [0.277, 0.297] | <0.001 | *** | 121,550 | Main Model |
| MoBa + SSB | Population | Birth Year | -0.013 | 0.001 | [-0.014, -0.011] | <0.001 | *** | 121,550 | Main Model |
| MoBa + SSB | Population | Birth Order | -0.048 | 0.002 | [-0.052, -0.045] | <0.001 | *** | 121,550 | Main Model |
| MoBa + SSB | Sibling | Intercept | 19.037 | 3.376 | [12.42, 25.66] | <0.001 | *** | 16,746 | Main Model |
| MoBa + SSB | Sibling | Bipolar PGS: Between-sibship | 0.025 | 0.009 | [0.007, 0.042] | 0.005 | ** | 16,746 | Main Model |
| MoBa + SSB | Sibling | Bipolar PGS: Within-sibship | 0.034 | 0.011 | [0.012, 0.055] | 0.001 | ** | 16,746 | Main Model |
| MoBa + SSB | Sibling | Sex | 0.281 | 0.014 | [0.254, 0.308] | <0.001 | *** | 16,746 | Main Model |
| MoBa + SSB | Sibling | Birth Year | -0.009 | 0.002 | [-0.013, -0.005] | <0.001 | *** | 16,746 | Main Model |
| MoBa + SSB | Sibling | Birth Order | -0.04 | 0.006 | [-0.052, -0.028] | <0.001 | *** | 16,746 | Main Model |
| UKB | Sibling | Intercept | | | | | | 12,284 | Main Model |
| UKB | Sibling | Bipolar PGS: Between-sibship | 0.029 | 0.011 | [0.007, 0.051] | <0.001 | *** | 12,284 | Main Model |
| UKB | Sibling | Bipolar PGS: Within-sibship | 0.015 | 0.015 | [-0.014, 0.044] | 0.3 | | 12,284 | Main Model |
| UKB | Sibling | Sex | | | | | | 12,284 | Main Model |
| UKB | Sibling | Birth Year | | | | | | 12,284 | Main Model |
| UKB | Sibling | Birth Order | | | | | | 12,284 | Main Model |

**Supplementary Table 7. Full regression results for sensitivity check for analysis 1 examining the association between bipolar disorder PGS and educational attainment (EA), adjusted for bipolar disorder diagnosis.**

*MoBa = the Norwegian Mother, Father and Child Cohort Study; SSB = Statistics Norway; UKB = UK Biobank; SE = Standard Error; CI = Confidence Interval.*
*p < .05. **p < .01. ***p < .001.

| Cohort | Model | Variable | Estimate | SE | 95% CI | p | Significance | N | Analysis |
|---|---|---|---|---|---|---|---|---|---|
| MoBa + SSB | Population | Intercept | 24.949 | 1.051 | [22.889, 27.009] | <0.001 | *** | 121,550 | Adjusted Model |
| MoBa + SSB | Population | Bipolar PGS | 0.028 | 0.003 | [0.022, 0.033] | <0.001 | *** | 121,550 | Adjusted Model |
| MoBa + SSB | Population | Bipolar Disorder Diagnosis | -0.228 | 0.026 | 0-0.279, -0.177] | <0.001 | *** | 121,550 | Adjusted Model |
| MoBa + SSB | Population | Sex | 0.288 | 0.005 | [0.278, 0.298] | <0.001 | *** | 121,550 | Adjusted Model |
| MoBa + SSB | Population | Birth Year | -0.012 | 0.001 | 0-0.014, -0.01] | <0.001 | *** | 121,550 | Adjusted Model |
| MoBa + SSB | Population | Birth Order | -0.048 | 0.002 | 0-0.052, -0.044] | <0.001 | *** | 121,550 | Adjusted Model |
| MoBa + SSB | Sibling | Intercept | 18.92 | 3.376 | [12.303, 25,537] | <0.001 | *** | 16,746 | Adjusted Model |
| MoBa + SSB | Sibling | Bipolar PGS: Between-sibship | 0.025 | 0.009 | [0.008, 0,043] | 0.0041 | ** | 16,746 | Adjusted Model |
| MoBa + SSB | Sibling | Bipolar PGS: Within-sibship | 0.034 | 0.011 | [0.013, 0,056] | 0.0012 | ** | 16,746 | Adjusted Model |
| MoBa + SSB | Sibling | Bipolar Disorder Diagnosis | -0.114 | 0.066 | [-0.244, 0,015] | 0.082 | | 16,746 | Adjusted Model |
| MoBa + SSB | Sibling | Sex | 0.281 | 0.014 | [0.254, 0,309] | <0.001 | *** | 16,746 | Adjusted Model |
| MoBa + SSB | Sibling | Birth Year | -0.009 | 0.002 | [-0.013, -0,005] | <0.001 | *** | 16,746 | Adjusted Model |
| MoBa + SSB | Sibling | Birth Order | -0.04 | 0.006 | [-0.052, -0,028] | <0.001 | *** | 16,746 | Adjusted Model |
| UKB | Sibling | Intercept | | | | | | 12,284 | Adjusted Model |
| UKB | Sibling | Bipolar PGS: Between-sibship | 0.029 | 0.011 | [0.007, 0.051] | <0.001 | *** | 12,284 | Adjusted Model |
| UKB | Sibling | Bipolar PGS: Within-sibship | 0.015 | 0.015 | [-0.014, 0.044] | 0.3 | | 12,284 | Adjusted Model |
| UKB | Sibling | Bipolar Disorder Diagnosis | | | | | | 12,284 | Adjusted Model |
| UKB | Sibling | Sex | | | | | | 12,284 | Adjusted Model |

| | | | | | | | | | |
|---|---|---|---|---|---|---|---|---|---|
| UKB | Sibling | Birth Year | | | | | | 12,284 | Adjusted Model |
| UKB | Sibling | Birth Order | | | | | | 12,284 | Adjusted Model |

**Supplementary Table 8. Full regression results for analysis 1 examining the association between bipolar disorder PGS and academic performance.**

*MoBa = the Norwegian Mother, Father and Child Cohort Study; SSB = Statistics Norway; SE = Standard Error; CI = Confidence Interval.*
**p < .05. **p < .01. ***p < .001.*

| Cohort | Age | Test | Model | Variable | Estimate | SE | 95% CI | p | Significance | N |
|---|---|---|---|---|---|---|---|---|---|---|
| MoBa + SSB | 10 | Total Score | Population | Intercept | -71.376 | 3.31 | [-77.863, -64.888] | <0.001 | | 70689 |
| MoBa + SSB | 10 | Total Score | Population | Bipolar Disorder PGS | 0.033 | 0.004 | [0.025, 0.041] | <0.001 | *** | 70689 |
| MoBa + SSB | 10 | Total Score | Population | Sex | -0.045 | 0.007 | [-0.058, -0.031] | <0.001 | *** | 70689 |
| MoBa + SSB | 10 | Total Score | Population | Birth Year | 0.036 | 0.002 | [0.032, 0.04] | <0.001 | *** | 70689 |
| MoBa + SSB | 10 | Total Score | Population | Birth Order | -0.048 | 0.005 | [-0.058, -0.038] | <0.001 | *** | 70689 |
| MoBa + SSB | 10 | Total Score | Sibling | Intercept | -65.902 | 8.052 | [-81.684, -50.12] | <0.001 | | 17677 |
| MoBa + SSB | 10 | Total Score | Sibling | Bipolar Disorder PGS: Between-sibship | 0.046 | 0.01 | [0.026, 0.065] | <0.001 | *** | 17677 |
| MoBa + SSB | 10 | Total Score | Sibling | Bipolar Disorder PGS: Within-sibship | 0.026 | 0.011 | [0.005, 0.048] | 0.0184 | * | 17677 |
| MoBa + SSB | 10 | Total Score | Sibling | Sex | -0.027 | 0.014 | [-0.054, 0.001] | 0.0622 | | 17677 |
| MoBa + SSB | 10 | Total Score | Sibling | Birth Year | 0.033 | 0.004 | [0.025, 0.041] | <0.001 | *** | 17677 |
| MoBa + SSB | 10 | Total Score | Sibling | Birth Order | -0.116 | 0.012 | [-0.14, -0.093] | <0.001 | *** | 17677 |
| MoBa + SSB | 10 | Math | Population | Intercept | -66.058 | 3.267 | [-72.461, -59.655] | <0.001 | | 72235 |
| MoBa + SSB | 10 | Math | Population | Bipolar Disorder PGS | 0.025 | 0.004 | [0.018, 0.033] | <0.001 | *** | 72235 |
| MoBa + SSB | 10 | Math | Population | Sex | -0.15 | 0.007 | [0.164, 0.137] | <0.001 | *** | 72235 |
| MoBa + SSB | 10 | Math | Population | Birth Year | 0.033 | 0.002 | [0.029, 0.037] | <0.001 | *** | 72235 |
| MoBa + SSB | 10 | Math | Population | Birth Order | -0.022 | 0.005 | [0.032, 0.012] | <0.001 | *** | 72235 |

| | | | | | | | | | | |
|---|---|---|---|---|---|---|---|---|---|---|
| MoBa + SSB | 10 | Math | Sibling | Intercept | -62.494 | 7.853 | [-77.886, -47.102] | <0.001 | | 18351 |
| MoBa + SSB | 10 | Math | Sibling | Bipolar Disorder PGS: Between-sibship | 0.034 | 0.01 | [0.015, 0.054] | <0.001 | *** | 18351 |
| MoBa + SSB | 10 | Math | Sibling | Bipolar Disorder PGS: Within-sibship | 0.015 | 0.011 | [-0.007, 0.036] | 0.1894 | | 18351 |
| MoBa + SSB | 10 | Math | Sibling | Sex | -0.136 | 0.014 | [-0.163, -0.108] | <0.001 | *** | 18351 |
| MoBa + SSB | 10 | Math | Sibling | Birth Year | 0.032 | 0.004 | [0.024, 0.039] | <0.001 | *** | 18351 |
| MoBa + SSB | 10 | Math | Sibling | Birth Order | -0.089 | 0.011 | [-0.111, -0.067] | <0.001 | *** | 18351 |
| MoBa + SSB | 10 | Reading | Population | Intercept | -60.53 | 3.284 | [-66.966, -54.093] | <0.001 | | 71306 |
| MoBa + SSB | 10 | Reading | Population | Bipolar Disorder PGS | 0.034 | 0.004 | [0.026, 0.042] | <0.001 | *** | 71306 |
| MoBa + SSB | 10 | Reading | Population | Sex | 0.127 | 0.007 | [0.114, 0.141] | <0.001 | *** | 71306 |
| MoBa + SSB | 10 | Reading | Population | Birth Year | 0.03 | 0.002 | [0.026, 0.034] | <0.001 | *** | 71306 |
| MoBa + SSB | 10 | Reading | Population | Birth Order | -0.075 | 0.005 | [-0.085, -0.065] | <0.001 | *** | 71306 |
| MoBa + SSB | 10 | Reading | Sibling | Intercept | -56.388 | 7.909 | [-71.889, -40.886] | <0.001 | | 17909 |
| MoBa + SSB | 10 | Reading | Sibling | Bipolar Disorder PGS: Between-sibship | 0.044 | 0.01 | [0.024, 0.063] | <0.001 | *** | 17909 |
| MoBa + SSB | 10 | Reading | Sibling | Bipolar Disorder PGS: Within-sibship | 0.035 | 0.012 | [0.012, 0.059] | 0.0024 | ** | 17909 |
| MoBa + SSB | 10 | Reading | Sibling | Sex | 0.142 | 0.014 | [0.115, 0.17] | <0.001 | *** | 17909 |
| MoBa + SSB | 10 | Reading | Sibling | Birth Year | 0.028 | 0.004 | [0.02, 0.036] | <0.001 | *** | 17909 |
| MoBa + SSB | 10 | Reading | Sibling | Birth Order | -0.138 | 0.011 | [-0.159, -0.116] | <0.001 | *** | 17909 |
| MoBa + SSB | 10 | English | Population | Intercept | -28.912 | 3.63 | [-36.026, -21.797] | <0.001 | | 68644 |
| MoBa + SSB | 10 | English | Population | Bipolar Disorder PGS | 0.043 | 0.004 | [0.036, 0.051] | <0.001 | *** | 68644 |
| MoBa + SSB | 10 | English | Population | Sex | -0.087 | 0.007 | [-0.1, -0.073] | <0.001 | *** | 68644 |
| MoBa + SSB | 10 | English | Population | Birth Year | 0.014 | 0.002 | [0.011, 0.018] | <0.001 | *** | 68644 |
| MoBa + SSB | 10 | English | Population | Birth Order | 0.019 | 0.005 | [0.01, 0.029] | <0.001 | *** | 68644 |
| MoBa + SSB | 10 | English | Sibling | Intercept | -53.678 | 8.784 | [-70.894, -36.461] | <0.001 | | 16835 |
| MoBa + SSB | 10 | English | Sibling | Bipolar Disorder PGS: Between-sibship | 0.056 | 0.01 | [0.037, 0.076] | <0.001 | *** | 16835 |
| MoBa + SSB | 10 | English | Sibling | Bipolar Disorder PGS: Within-sibship | 0.034 | 0.012 | [0.011, 0.058] | 0.004 | ** | 16835 |
| MoBa + SSB | 10 | English | Sibling | Sex | -0.074 | 0.015 | [-0.104, -0.045] | <0.001 | *** | 16835 |

| | | | | | | | | | | |
|---|---|---|---|---|---|---|---|---|---|---|
| MoBa + SSB | 10 | English | Sibling | Birth Year | 0.027 | 0.004 | [0.019, 0.035] | <0.001 | *** | 16835 |
| MoBa + SSB | 10 | English | Sibling | Birth Order | -0.038 | 0.012 | [-0.062, -0.014] | 0.0017 | ** | 16835 |
| MoBa + SSB | 13 | Total Score | Population | Intercept | -67.512 | 3.261 | [-73.904, -61.121] | <0.001 | | 71320 |
| MoBa + SSB | 13 | Total Score | Population | Bipolar Disorder PGS | 0.028 | 0.004 | [0.02, 0.036] | <0.001 | *** | 71320 |
| MoBa + SSB | 13 | Total Score | Population | Sex | 0.006 | 0.007 | [-0.008, 0.019] | 0.4219 | | 71320 |
| MoBa + SSB | 13 | Total Score | Population | Birth Year | 0.034 | 0.002 | [0.03, 0.038] | <0.001 | *** | 71320 |
| MoBa + SSB | 13 | Total Score | Population | Birth Order | -0.048 | 0.005 | [-0.057, -0.038] | <0.001 | *** | 71320 |
| MoBa + SSB | 13 | Total Score | Sibling | Intercept | -50.092 | 7.952 | [-65.678, -34.506] | <0.001 | | 17916 |
| MoBa + SSB | 13 | Total Score | Sibling | Bipolar Disorder PGS: Between-sibship | 0.037 | 0.01 | [0.017, 0.057] | <0.001 | *** | 17916 |
| MoBa + SSB | 13 | Total Score | Sibling | Bipolar Disorder PGS: Within-sibship | 0.011 | 0.01 | [-0.009, 0.03] | 0.3011 | | 17916 |
| MoBa + SSB | 13 | Total Score | Sibling | Sex | 0.015 | 0.014 | [-0.012, 0.042] | 0.2855 | | 17916 |
| MoBa + SSB | 13 | Total Score | Sibling | Birth Year | 0.025 | 0.004 | [0.017, 0.033] | <0.001 | *** | 17916 |
| MoBa + SSB | 13 | Total Score | Sibling | Birth Order | -0.118 | 0.011 | [-0.139, -0.096] | <0.001 | *** | 17916 |
| MoBa + SSB | 13 | Math | Population | Intercept | -57.482 | 3.28 | [-63.911, -51.054] | <0.001 | | 72192 |
| MoBa + SSB | 13 | Math | Population | Bipolar Disorder PGS | 0.023 | 0.004 | [0.015, 0.031] | <0.001 | *** | 72192 |
| MoBa + SSB | 13 | Math | Population | Sex | -0.191 | 0.007 | [-0.204, -0.177] | <0.001 | *** | 72192 |
| MoBa + SSB | 13 | Math | Population | Birth Year | 0.029 | 0.002 | [0.025, 0.033] | <0.001 | *** | 72192 |
| MoBa + SSB | 13 | Math | Population | Birth Order | -0.024 | 0.005 | [-0.034, -0.014] | <0.001 | *** | 72192 |
| MoBa + SSB | 13 | Math | Sibling | Intercept | -45.763 | 7.899 | [-61.245, -30.281] | <0.001 | | 18277 |
| MoBa + SSB | 13 | Math | Sibling | Bipolar Disorder PGS: Between-sibship | 0.034 | 0.01 | [0.015, 0.054] | <0.001 | *** | 18277 |
| MoBa + SSB | 13 | Math | Sibling | Bipolar Disorder PGS: Within-sibship | 0.005 | 0.011 | [-0.016, 0.027] | 0.6226 | | 18277 |
| MoBa + SSB | 13 | Math | Sibling | Sex | -0.18 | 0.014 | [-0.207, -0.153] | <0.001 | *** | 18277 |
| MoBa + SSB | 13 | Math | Sibling | Birth Year | 0.023 | 0.004 | [0.015, 0.031] | <0.001 | *** | 18277 |
| MoBa + SSB | 13 | Math | Sibling | Birth Order | -0.084 | 0.011 | [-0.106, -0.063] | <0.001 | *** | 18277 |
| MoBa + SSB | 13 | Reading | Population | Intercept | -64.833 | 3.186 | [-71.078, -58.589] | <0.001 | | 72125 |

| | | | | | | | | | | |
|---|---|---|---|---|---|---|---|---|---|---|
| MoBa + SSB | 13 | Reading | Population | Bipolar Disorder PGS | 0.028 | 0.004 | [0.02, 0.035] | <0.001 | *** | 72125 |
| MoBa + SSB | 13 | Reading | Population | Sex | 0.243 | 0.007 | [0.229, 0.256] | <0.001 | *** | 72125 |
| MoBa + SSB | 13 | Reading | Population | Birth Year | 0.032 | 0.002 | [0.028, 0.036] | <0.001 | *** | 72125 |
| MoBa + SSB | 13 | Reading | Population | Birth Order | -0.067 | 0.005 | [-0.077, -0.058] | <0.001 | *** | 72125 |
| MoBa + SSB | 13 | Reading | Sibling | Intercept | -46.545 | 7.71 | [-61.656, -31.433] | <0.001 | | 18245 |
| MoBa + SSB | 13 | Reading | Sibling | Bipolar Disorder PGS: Between-sibship | 0.033 | 0.01 | [0.014, 0.053] | <0.001 | *** | 18245 |
| MoBa + SSB | 13 | Reading | Sibling | Bipolar Disorder PGS: Within-sibship | 0.018 | 0.011 | [-0.004, 0.04] | 0.0947 | | 18245 |
| MoBa + SSB | 13 | Reading | Sibling | Sex | 0.249 | 0.014 | [0.222, 0.277] | <0.001 | *** | 18245 |
| MoBa + SSB | 13 | Reading | Sibling | Birth Year | 0.023 | 0.004 | [0.015, 0.031] | <0.001 | *** | 18245 |
| MoBa + SSB | 13 | Reading | Sibling | Birth Order | -0.14 | 0.011 | [-0.162, -0.119] | <0.001 | *** | 18245 |
| MoBa + SSB | 13 | English | Population | Intercept | -45.428 | 3.314 | [-51.923, -38.932] | <0.001 | | 71849 |
| MoBa + SSB | 13 | English | Population | Bipolar Disorder PGS | 0.046 | 0.004 | [0.038, 0.054] | <0.001 | *** | 71849 |
| MoBa + SSB | 13 | English | Population | Sex | -0.073 | 0.007 | [-0.086, -0.059] | <0.001 | *** | 71849 |
| MoBa + SSB | 13 | English | Population | Birth Year | 0.023 | 0.002 | [0.019, 0.027] | <0.001 | *** | 71849 |
| MoBa + SSB | 13 | English | Population | Birth Order | -0.007 | 0.005 | [-0.017, 0.003] | 0.1245 | | 71849 |
| MoBa + SSB | 13 | English | Sibling | Intercept | -52.548 | 8.12 | [-68.464, -36.633] | <0.001 | | 18163 |
| MoBa + SSB | 13 | English | Sibling | Bipolar Disorder PGS: Between-sibship | 0.063 | 0.01 | [0.043, 0.082] | <0.001 | *** | 18163 |
| MoBa + SSB | 13 | English | Sibling | Bipolar Disorder PGS: Within-sibship | 0.02 | 0.011 | [-0.002, 0.041] | 0.0762 | | 18163 |
| MoBa + SSB | 13 | English | Sibling | Sex | -0.061 | 0.015 | [-0.091, -0.032] | <0.001 | *** | 18163 |
| MoBa + SSB | 13 | English | Sibling | Birth Year | 0.026 | 0.004 | [0.019, 0.034] | <0.001 | *** | 18163 |
| MoBa + SSB | 13 | English | Sibling | Birth Order | -0.058 | 0.012 | [-0.082, -0.035] | <0.001 | *** | 18163 |
| MoBa + SSB | 14 | Total Score | Population | Intercept | -58.388 | 3.204 | [-64.668, -52.108] | <0.001 | | 69779 |
| MoBa + SSB | 14 | Total Score | Population | Bipolar Disorder PGS | 0.022 | 0.004 | [0.014, 0.03] | <0.001 | *** | 69779 |
| MoBa + SSB | 14 | Total Score | Population | Sex | 0.035 | 0.007 | [0.022, 0.049] | <0.001 | *** | 69779 |
| MoBa + SSB | 14 | Total Score | Population | Birth Year | 0.029 | 0.002 | [0.025, 0.033] | <0.001 | *** | 69779 |
| MoBa + SSB | 14 | Total Score | Population | Birth Order | -0.042 | 0.004 | [-0.049, -0.034] | <0.001 | *** | 69779 |

| | | | | | | | | | | |
|---|---|---|---|---|---|---|---|---|---|---|
| MoBa + SSB | 14 | Total Score | Sibling | Intercept | -45.986 | 7.821 | [-61.315, -30.657] | <0.001 | | 17172 |
| MoBa + SSB | 14 | Total Score | Sibling | Bipolar Disorder PGS: Between-sibship | 0.037 | 0.01 | [0.017, 0.056] | <0.001 | *** | 17172 |
| MoBa + SSB | 14 | Total Score | Sibling | Bipolar Disorder PGS: Within-sibship | 0.02 | 0.011 | [-0.002, 0.042] | 0.0602 | | 17172 |
| MoBa + SSB | 14 | Total Score | Sibling | Sex | 0.039 | 0.014 | [0.012, 0.067] | 0.0045 | ** | 17172 |
| MoBa + SSB | 14 | Total Score | Sibling | Birth Year | 0.023 | 0.004 | [0.015, 0.031] | <0.001 | *** | 17172 |
| MoBa + SSB | 14 | Total Score | Sibling | Birth Order | -0.115 | 0.011 | [-0.136, -0.093] | <0.001 | *** | 17172 |
| MoBa + SSB | 14 | Math | Population | Intercept | -52.049 | 3.234 | [-58.388, -45,711] | <0.001 | | 70718 |
| MoBa + SSB | 14 | Math | Population | Bipolar Disorder PGS | 0.016 | 0.004 | [0.008, -0,024] | <0.001 | *** | 70718 |
| MoBa + SSB | 14 | Math | Population | Sex | -0.171 | 0.007 | [-0.185, -0,158] | <0.001 | *** | 70718 |
| MoBa + SSB | 14 | Math | Population | Birth Year | 0.026 | 0.002 | [0.022, -0,03] | <0.001 | *** | 70718 |
| MoBa + SSB | 14 | Math | Population | Birth Order | -0.02 | 0.004 | [-0.028, -0,013] | <0.001 | *** | 70718 |
| MoBa + SSB | 14 | Math | Sibling | Intercept | -38.803 | 7.746 | [-53.985, -23.621] | <0.001 | | 17635 |
| MoBa + SSB | 14 | Math | Sibling | Bipolar Disorder PGS: Between-sibship | 0.032 | 0.01 | [0.013, 0.052] | <0.001 | *** | 17635 |
| MoBa + SSB | 14 | Math | Sibling | Bipolar Disorder PGS: Within-sibship | 0.009 | 0.011 | [-0.012, 0.031] | 0.382 | | 17635 |
| MoBa + SSB | 14 | Math | Sibling | Sex | -0.157 | 0.014 | [-0.184, -0.129] | <0.001 | *** | 17635 |
| MoBa + SSB | 14 | Math | Sibling | Birth Year | 0.02 | 0.004 | [0.012, 0.028] | <0.001 | *** | 17635 |
| MoBa + SSB | 14 | Math | Sibling | Birth Order | -0.083 | 0.011 | [-0.104, -0.061] | <0.001 | *** | 17635 |
| MoBa + SSB | 14 | Reading | Population | Intercept | -53.77 | 3.117 | [-59.879, -47.661] | <0.001 | | 71017 |
| MoBa + SSB | 14 | Reading | Population | Bipolar Disorder PGS | 0.023 | 0.003 | [0.017, 0.029] | <0.001 | *** | 71017 |
| MoBa + SSB | 14 | Reading | Population | Sex | 0.275 | 0.007 | [0.261, 0.289] | <0.001 | *** | 71017 |
| MoBa + SSB | 14 | Reading | Population | Birth Year | 0.027 | 0.002 | [0.023, 0.031] | <0.001 | *** | 71017 |
| MoBa + SSB | 14 | Reading | Population | Birth Order | -0.059 | 0.004 | [-0.067, -0.052] | <0.001 | *** | 71017 |
| MoBa + SSB | 14 | Reading | Sibling | Intercept | -42.724 | 7.592 | [-57.604, -27.844] | <0.001 | | 17713 |
| MoBa + SSB | 14 | Reading | Sibling | Bipolar Disorder PGS: Between-sibship | 0.027 | 0.009 | [0.01, 0.045] | 0.0027 | ** | 17713 |
| MoBa + SSB | 14 | Reading | Sibling | Bipolar Disorder PGS: Within-sibship | 0.024 | 0.011 | [0.003, 0.046] | 0.0226 | * | 17713 |

| | | | | | | | | | | |
|---|---|---|---|---|---|---|---|---|---|---|
| MoBa + SSB | 14 | Reading | Sibling | Sex | 0.276 | 0.013 | [0.25, 0.301] | <0.001 | *** | 17713 |
| MoBa + SSB | 14 | Reading | Sibling | Birth Year | 0.021 | 0.004 | [0.014, 0.029] | <0.001 | *** | 17713 |
| MoBa + SSB | 14 | Reading | Sibling | Birth Order | -0.133 | 0.011 | [-0.155, -0.111] | <0.001 | *** | 17713 |

**Supplementary Table 9. Full regression results for analysis 2 examining the association between academic performance and bipolar disorder diagnosis.**

*SSB = Statistics Norway; CBS = Statistics Netherlands; SE = Standard Error; CI = Confidence Interval.*
**p < .05. **p < .01. ***p < .001.*

| Cohort | Age | Test | Model | Variable | Estimate | SE | HR | 95% CI | p | Significance | N | Analysis |
|---|---|---|---|---|---|---|---|---|---|---|---|---|
| SSB | 10 | Total Score | Population | Age 10 Total Score | 0.039 | 0.019 | 1.04 | [1.002, 1.079] | 0.0449 | * | 768505 | Main Model |
| SSB | 10 | Total Score | Population | Sex | 0.863 | 0.041 | 2.37 | [2.187, 2.569] | <0.001 | *** | 768505 | Main Model |
| SSB | 10 | Total Score | Population | Birth Year | -0.14 | 0.006 | 0.869 | [0.859, 0.88] | <0.001 | *** | 768505 | Main Model |
| SSB | 10 | Total Score | Population | Birth Order | 0.121 | 0.017 | 1.129 | [1.092, 1.167] | <0.001 | *** | 768505 | Main Model |
| SSB | 10 | Total Score | Sibling | Age 10 Total Score: Between-sibship | -0.001 | 0.035 | 0.999 | [0.933, 1.07] | 0.9788 | | 343898 | Main Model |
| SSB | 10 | Total Score | Sibling | Age 10 Total Score: Within-sibship | 0.109 | 0.048 | 1.115 | [1.015, 1.225] | 0.0241 | * | 343898 | Main Model |
| SSB | 10 | Total Score | Sibling | Sex | 0.824 | 0.06 | 2.28 | [2.027, 2.564] | <0.001 | *** | 343898 | Main Model |
| SSB | 10 | Total Score | Sibling | Birth Year | -0.143 | 0.009 | 0.867 | [0.852, 0.882] | <0.001 | *** | 343898 | Main Model |
| SSB | 10 | Total Score | Sibling | Birth Order | 0.004 | 0.032 | 1.004 | [0.943, 1.069] | 0.9115 | | 343898 | Main Model |
| SSB | 10 | Math | Population | Age 10 Math Score | -0.022 | 0.019 | 0.978 | [0.942, 1.015] | 0.2396 | | 791751 | Main Model |
| SSB | 10 | Math | Population | Sex | 0.864 | 0.041 | 2.373 | [2.189, 2.571] | <0.001 | *** | 791751 | Main Model |
| SSB | 10 | Math | Population | Birth Year | -0.14 | 0.006 | 0.869 | [0.859, 0.88] | <0.001 | *** | 791751 | Main Model |
| SSB | 10 | Math | Population | Birth Order | 0.118 | 0.017 | 1.125 | [1.088, 1.163] | <0.001 | *** | 791751 | Main Model |
| SSB | 10 | Math | Sibling | Age 10 Math Score: Between-sibship | -0.089 | 0.035 | 0.915 | [0.854, 0.98] | 0.0106 | * | 359395 | Main Model |
| SSB | 10 | Math | Sibling | Age 10 Math Score: Within-sibship | 0.051 | 0.046 | 1.052 | [0.962, 1.152] | 0.2655 | | 359395 | Main Model |
| SSB | 10 | Math | Sibling | Sex | 0.812 | 0.058 | 2.252 | [2.01, 2.524] | <0.001 | *** | 359395 | Main Model |

| | | | | | | | | | | | | |
|---|---|---|---|---|---|---|---|---|---|---|---|---|
| SSB | 10 | Math | Sibling | Birth Year | -0.142 | 0.009 | 0.868 | [0.852, 0.883] | <0.001 | *** | 359395 | Main Model |
| SSB | 10 | Math | Sibling | Birth Order | -0.002 | 0.031 | 0.998 | [0.939, 1.061] | 0.9407 | | 359395 | Main Model |
| SSB | 10 | Reading | Population | Age 10 Reading Score | 0.126 | 0.02 | 1.134 | [1.091, 1.18] | <0.001 | *** | 779890 | Main Model |
| SSB | 10 | Reading | Population | Sex | 0.848 | 0.041 | 2.335 | [2.155, 2.53] | <0.001 | *** | 779890 | Main Model |
| SSB | 10 | Reading | Population | Birth Year | -0.14 | 0.006 | 0.869 | [0.859, 0.88] | <0.001 | *** | 779890 | Main Model |
| SSB | 10 | Reading | Population | Birth Order | 0.124 | 0.017 | 1.132 | [1.095, 1.17] | <0.001 | *** | 779890 | Main Model |
| SSB | 10 | Reading | Sibling | Age 10 Reading Score: Between-sibship | 0.128 | 0.036 | 1.137 | [1.059, 1.22] | <0.001 | *** | 351087 | Main Model |
| SSB | 10 | Reading | Sibling | Age 10 Reading Score: Within-sibship | 0.154 | 0.046 | 1.166 | [1.066, 1.277] | <0.001 | *** | 351087 | Main Model |
| SSB | 10 | Reading | Sibling | Sex | 0.818 | 0.059 | 2.266 | [2.019, 2.544] | <0.001 | *** | 351087 | Main Model |
| SSB | 10 | Reading | Sibling | Birth Year | -0.145 | 0.009 | 0.865 | [0.85, 0.88] | <0.001 | *** | 351087 | Main Model |
| SSB | 10 | Reading | Sibling | Birth Order | 0.004 | 0.032 | 1.004 | [0.943, 1.069] | 0.8932 | | 351087 | Main Model |
| SSB | 10 | English | Population | Age 10 English Score | 0.13 | 0.021 | 1.139 | [1.093, 1.187] | <0.001 | *** | 733986 | Main Model |
| SSB | 10 | English | Population | Sex | 0.868 | 0.043 | 2.382 | [2.19, 2.592] | <0.001 | *** | 733986 | Main Model |
| SSB | 10 | English | Population | Birth Year | -0.14 | 0.006 | 0.869 | [0.859, 0.88] | <0.001 | *** | 733986 | Main Model |
| SSB | 10 | English | Population | Birth Order | 0.125 | 0.018 | 1.133 | [1.094, 1.174] | <0.001 | *** | 733986 | Main Model |
| SSB | 10 | English | Sibling | Age 10 English Score: Between-sibship | 0.152 | 0.041 | 1.164 | [1.074, 1.262] | <0.001 | *** | 313825 | Main Model |
| SSB | 10 | English | Sibling | Age 10 English Score: Within-sibship | 0.077 | 0.051 | 1.08 | [0.977, 1.194] | 0.1269 | | 313825 | Main Model |
| SSB | 10 | English | Sibling | Sex | 0.806 | 0.065 | 2.239 | [1.971, 2.543] | <0.001 | *** | 313825 | Main Model |
| SSB | 10 | English | Sibling | Birth Year | -0.147 | 0.009 | 0.863 | [0.848, 0.879] | <0.001 | *** | 313825 | Main Model |
| SSB | 10 | English | Sibling | Birth Order | 0.036 | 0.033 | 1.037 | [0.972, 1.106] | 0.2754 | | 313825 | Main Model |
| CBS | 12 | Total Score | Population | Age 12 Total Score | 0.23 | 0.057 | 1.259 | [1.126, 1.407] | <0.001 | *** | 1144287 | Main Model |
| CBS | 12 | Total Score | Population | Sex | 0.182 | 0.104 | 1.2 | [0.978, 1.471] | 0.0807 | | 1144287 | Main Model |
| CBS | 12 | Total Score | Population | Birth Year | -0.082 | 0.031 | 0.921 | [0.867, 0.979] | 0.0084 | ** | 1144287 | Main Model |
| CBS | 12 | Total Score | Population | Birth Order | -0.127 | 0.083 | 0.881 | [0.749, 1.036] | 0.1227 | | 1144287 | Main Model |

| | | | | | | | | | | | | |
|---|---|---|---|---|---|---|---|---|---|---|---|---|
| CBS | 12 | Total Score | Sibling | Age 12 Total Score: Between-sibship | 0.186 | 0.098 | 1.204 | [0.994, 1.459] | 0.0570 | | 619796 | Main Model |
| CBS | 12 | Total Score | Sibling | Age 12 Total Score: Within-sibship | 0.467 | 0.161 | 1.595 | [1.164, 2.187] | 0.0038 | ** | 619796 | Main Model |
| CBS | 12 | Total Score | Sibling | Sex | 0.251 | 0.155 | 1.285 | [0.949, 1.742] | 0.1052 | | 619796 | Main Model |
| CBS | 12 | Total Score | Sibling | Birth Year | -0.055 | 0.047 | 0.946 | [0.863, 1.038] | 0.2419 | | 619796 | Main Model |
| CBS | 12 | Total Score | Sibling | Birth Order | -0.077 | 0.13 | 0.926 | [0.718, 1.195] | 0.5511 | | 619796 | Main Model |
| CBS | 12 | Math | Population | Age 12 Math Score | 0.163 | 0.056 | 1.177 | [1.055, 1.314] | 0.0036 | ** | 1144196 | Main Model |
| CBS | 12 | Math | Population | Sex | 0.227 | 0.107 | 1.255 | [1.017, 1.548] | 0.0347 | * | 1144196 | Main Model |
| CBS | 12 | Math | Population | Birth Year | -0.078 | 0.031 | 0.925 | [0.87, 0.983] | 0.0108 | * | 1144196 | Main Model |
| CBS | 12 | Math | Population | Birth Order | -0.138 | 0.083 | 0.871 | [0.74, 1.025] | 0.0951 | | 1144196 | Main Model |
| CBS | 12 | Math | Sibling | Age 12 Math Score: Between-sibship | 0.134 | 0.102 | 1.143 | [0.936, 1.396] | 0.1909 | | 619735 | Main Model |
| CBS | 12 | Math | Sibling | Age 12 Math Score: Within-sibship | 0.31 | 0.152 | 1.363 | [1.012, 1.837] | 0.0418 | * | 619735 | Main Model |
| CBS | 12 | Math | Sibling | Sex | 0.308 | 0.16 | 1.361 | [0.994, 1.862] | 0.0544 | | 619735 | Main Model |
| CBS | 12 | Math | Sibling | Birth Year | -0.054 | 0.047 | 0.947 | [0.864, 1.039] | 0.2504 | | 619735 | Main Model |
| CBS | 12 | Math | Sibling | Birth Order | -0.099 | 0.131 | 0.906 | [0.701, 1.171] | 0.4513 | | 619735 | Main Model |
| CBS | 12 | Reading | Population | Age 12 Reading Score | 0.138 | 0.059 | 1.148 | [1.023, 1.289] | 0.0193 | * | 1142246 | Main Model |
| CBS | 12 | Reading | Population | Sex | 0.173 | 0.104 | 1.189 | [0.97, 1.458] | 0.0948 | | 1142246 | Main Model |
| CBS | 12 | Reading | Population | Birth Year | -0.078 | 0.031 | 0.925 | [0.87, 0.983] | 0.0119 | * | 1142246 | Main Model |
| CBS | 12 | Reading | Population | Birth Order | -0.128 | 0.082 | 0.88 | [0.749, 1.033] | 0.1192 | | 1142246 | Main Model |
| CBS | 12 | Reading | Sibling | Age 12 Reading Score: Between-sibship | 0.075 | 0.103 | 1.078 | [0.881, 1.319] | 0.4662 | | 618519 | Main Model |
| CBS | 12 | Reading | Sibling | Age 12 Reading Score: Within-sibship | 0.363 | 0.15 | 1.438 | [1.071, 1.929] | 0.0157 | * | 618519 | Main Model |
| CBS | 12 | Reading | Sibling | Sex | 0.24 | 0.154 | 1.271 | [0.94, 1.719] | 0.1196 | | 618519 | Main Model |
| CBS | 12 | Reading | Sibling | Birth Year | -0.05 | 0.047 | 0.951 | [0.868, 1.043] | 0.2850 | | 618519 | Main Model |
| CBS | 12 | Reading | Sibling | Birth Order | -0.086 | 0.128 | 0.918 | [0.714, 1.179] | 0.5035 | | 618519 | Main Model |
| CBS | 12 | Language | Population | Age 12 Language Score | 0.284 | 0.058 | 1.328 | [1.186, 1.488] | <0.001 | *** | 1144196 | Main Model |
| CBS | 12 | Language | Population | Sex | 0.117 | 0.103 | 1.124 | [0.919, 1.376] | 0.2556 | | 1144196 | Main Model |

| | | | | | | | | | | | | |
|---|---|---|---|---|---|---|---|---|---|---|---|---|
| CBS | 12 | Language | Population | Birth Year | -0.08 | 0.031 | 0.923 | [0.869, 0.981] | 0.0098 | ** | 1144196 | Main Model |
| CBS | 12 | Language | Population | Birth Order | -0.118 | 0.082 | 0.889 | [0.757, 1.044] | 0.1514 | | 1144196 | Main Model |
| CBS | 12 | Language | Sibling | Age 12 Language Score: Between-sibship | 0.258 | 0.095 | 1.294 | [1.074, 1.559] | 0.0069 | ** | 619735 | Main Model |
| CBS | 12 | Language | Sibling | Age 12 Language Score: Within-sibship | 0.521 | 0.159 | 1.684 | [1.233, 2.299] | 0.0010 | ** | 619735 | Main Model |
| CBS | 12 | Language | Sibling | Sex | 0.166 | 0.153 | 1.181 | [0.875, 1.593] | 0.2784 | | 619735 | Main Model |
| CBS | 12 | Language | Sibling | Birth Year | -0.053 | 0.047 | 0.948 | [0.865, 1.04] | 0.2577 | | 619735 | Main Model |
| CBS | 12 | Language | Sibling | Birth Order | -0.063 | 0.129 | 0.939 | [0.729, 1.209] | 0.6243 | | 619735 | Main Model |
| SSB | 13 | Total Score | Population | Age 13 Total Score | -0.009 | 0.015 | 0.991 | [0.962, 1.021] | 0.5726 | | 949575 | Main Model |
| SSB | 13 | Total Score | Population | Sex | 0.773 | 0.032 | 2.166 | [2.035, 2.306] | <0.001 | *** | 949575 | Main Model |
| SSB | 13 | Total Score | Population | Birth Year | -0.094 | 0.004 | 0.91 | [0.903, 0.917] | <0.001 | *** | 949575 | Main Model |
| SSB | 13 | Total Score | Population | Birth Order | 0.102 | 0.014 | 1.107 | [1.077, 1.138] | <0.001 | *** | 949575 | Main Model |
| SSB | 13 | Total Score | Sibling | Age 13 Total Score: Between-sibship | -0.038 | 0.025 | 0.963 | [0.917, 1.011] | 0.1341 | | 449103 | Main Model |
| SSB | 13 | Total Score | Sibling | Age 13 Total Score: Within-sibship | 0.007 | 0.039 | 1.007 | [0.933, 1.087] | 0.8643 | | 449103 | Main Model |
| SSB | 13 | Total Score | Sibling | Sex | 0.772 | 0.045 | 2.164 | [1.981, 2.364] | <0.001 | *** | 449103 | Main Model |
| SSB | 13 | Total Score | Sibling | Birth Year | -0.097 | 0.006 | 0.908 | [0.897, 0.918] | <0.001 | *** | 449103 | Main Model |
| SSB | 13 | Total Score | Sibling | Birth Order | -0.018 | 0.024 | 0.982 | [0.937, 1.029] | 0.4487 | | 449103 | Main Model |
| SSB | 13 | Math | Population | Age 13 Math Score | -0.078 | 0.015 | 0.925 | [0.898, 0.953] | <0.001 | *** | 976455 | Main Model |
| SSB | 13 | Math | Population | Sex | 0.765 | 0.031 | 2.149 | [2.022, 2.284] | <0.001 | *** | 976455 | Main Model |
| SSB | 13 | Math | Population | Birth Year | -0.095 | 0.004 | 0.909 | [0.902, 0.917] | <0.001 | *** | 976455 | Main Model |
| SSB | 13 | Math | Population | Birth Order | 0.101 | 0.013 | 1.106 | [1.078, 1.135] | <0.001 | *** | 976455 | Main Model |
| SSB | 13 | Math | Sibling | Age 13 Math Score: Between-sibship | -0.13 | 0.025 | 0.878 | [0.836, 0.922] | <0.001 | *** | 466907 | Main Model |
| SSB | 13 | Math | Sibling | Age 13 Math Score: Within-sibship | -0.031 | 0.037 | 0.969 | [0.902, 1.042] | 0.3975 | | 466907 | Main Model |
| SSB | 13 | Math | Sibling | Sex | 0.765 | 0.043 | 2.149 | [1.975, 2.338] | <0.001 | *** | 466907 | Main Model |
| SSB | 13 | Math | Sibling | Birth Year | -0.1 | 0.006 | 0.905 | [0.894, 0.916] | <0.001 | *** | 466907 | Main Model |
| SSB | 13 | Math | Sibling | Birth Order | -0.02 | 0.023 | 0.98 | [0.937, 1.025] | 0.3849 | | 466907 | Main Model |

| | | | | | | | | | | | | |
|---|---|---|---|---|---|---|---|---|---|---|---|---|
| SSB | 13 | Reading | Population | Age 13 Reading Score | 0.087 | 0.015 | 1.091 | [1.059, 1.123] | <0.001 | *** | 969899 | Main Model |
| SSB | 13 | Reading | Population | Sex | 0.731 | 0.032 | 2.077 | [1.951, 0.917] | <0.001 | *** | 969899 | Main Model |
| SSB | 13 | Reading | Population | Birth Year | -0.094 | 0.004 | 0.91 | [0.903, 0.917] | <0.001 | *** | 969899 | Main Model |
| SSB | 13 | Reading | Population | Birth Order | 0.105 | 0.014 | 1.111 | [1.081, 1.142] | <0.001 | *** | 969899 | Main Model |
| SSB | 13 | Reading | Sibling | Age 13 Reading Score: Between-sibship | 0.076 | 0.026 | 1.079 | [1.025, 1.135] | 0.0031 | ** | 462351 | Main Model |
| SSB | 13 | Reading | Sibling | Age 13 Reading Score: Within-sibship | 0.057 | 0.037 | 1.059 | [0.985, 1.138] | 0.1290 | | 462351 | Main Model |
| SSB | 13 | Reading | Sibling | Sex | 0.733 | 0.044 | 2.081 | [1.909, 2.269] | <0.001 | *** | 462351 | Main Model |
| SSB | 13 | Reading | Sibling | Birth Year | -0.098 | 0.006 | 0.907 | [0.896, 0.917] | <0.001 | *** | 462351 | Main Model |
| SSB | 13 | Reading | Sibling | Birth Order | -0.01 | 0.023 | 0.99 | [0.946, 1.036] | 0.6628 | | 462351 | Main Model |
| SSB | 13 | English | Population | Age 13 English Score | 0.186 | 0.016 | 1.204 | [1.167, 1.243] | <0.001 | *** | 971859 | Main Model |
| SSB | 13 | English | Population | Sex | 0.769 | 0.031 | 2.158 | [2.03, 2.293] | <0.001 | *** | 971859 | Main Model |
| SSB | 13 | English | Population | Birth Year | -0.094 | 0.004 | 0.91 | [0.903, 0.917] | <0.001 | *** | 971859 | Main Model |
| SSB | 13 | English | Population | Birth Order | 0.103 | 0.013 | 1.108 | [1.081, 1.137] | <0.001 | *** | 971859 | Main Model |
| SSB | 13 | English | Sibling | Age 13 English Score: Between-sibship | 0.219 | 0.027 | 1.245 | [1.181, 1.312] | <0.001 | *** | 462721 | Main Model |
| SSB | 13 | English | Sibling | Age 13 English Score: Within-sibship | 0.184 | 0.038 | 1.202 | [1.116, 1.295] | <0.001 | *** | 462721 | Main Model |
| SSB | 13 | English | Sibling | Sex | 0.775 | 0.043 | 2.171 | [1.995, 2.361] | <0.001 | *** | 462721 | Main Model |
| SSB | 13 | English | Sibling | Birth Year | -0.101 | 0.006 | 0.904 | [0.893, 0.915] | <0.001 | *** | 462721 | Main Model |
| SSB | 13 | English | Sibling | Birth Order | -0.006 | 0.022 | 0.994 | [0.952, 1.038] | 0.7948 | | 462721 | Main Model |
| SSB | 14 | Total Score | Population | Age 14 Total Score | -0.072 | 0.018 | 0.931 | [0.898, 0.964] | <0.001 | *** | 833916 | Main Model |
| SSB | 14 | Total Score | Population | Sex | 0.822 | 0.038 | 2.275 | [2.112, 2.451] | <0.001 | *** | 833916 | Main Model |
| SSB | 14 | Total Score | Population | Birth Year | -0.124 | 0.005 | 0.883 | [0.875, 0.892] | <0.001 | *** | 833916 | Main Model |
| SSB | 14 | Total Score | Population | Birth Order | 0.104 | 0.016 | 1.11 | [1.075, 1.145] | <0.001 | *** | 833916 | Main Model |
| SSB | 14 | Total Score | Sibling | Age 14 Total Score: Between-sibship | -0.093 | 0.031 | 0.911 | [0.857, 0.968] | 0.0029 | ** | 382566 | Main Model |
| SSB | 14 | Total Score | Sibling | Age 14 Total Score: Within-sibship | -0.047 | 0.047 | 0.954 | [0.87, 1.046] | 0.3257 | | 382566 | Main Model |

| | | | | | | | | | | | | |
|---|---|---|---|---|---|---|---|---|---|---|---|---|
| SSB | 14 | Total Score | Sibling | Sex | 0.799 | 0.055 | 2.223 | [1.996, 2.476] | <0.001 | *** | 382566 | Main Model |
| SSB | 14 | Total Score | Sibling | Birth Year | -0.131 | 0.008 | 0.877 | [0.864, 0.891] | <0.001 | *** | 382566 | Main Model |
| SSB | 14 | Total Score | Sibling | Birth Order | 0.011 | 0.028 | 1.011 | [0.957, 1.068] | 0.6967 | | 382566 | Main Model |
| SSB | 14 | Math | Population | Age 14 Math Score | -0.133 | 0.018 | 0.875 | [0.845, 0.907] | <0.001 | *** | 755298 | Main Model |
| SSB | 14 | Math | Population | Sex | 0.804 | 0.037 | 2.234 | [2.078, 2.403] | <0.001 | *** | 755298 | Main Model |
| SSB | 14 | Math | Population | Birth Year | -0.122 | 0.005 | 0.885 | [0.877, 0.894] | <0.001 | *** | 755298 | Main Model |
| SSB | 14 | Math | Population | Birth Order | 0.099 | 0.016 | 1.104 | [1.07, 1.139] | <0.001 | *** | 755298 | Main Model |
| SSB | 14 | Math | Sibling | Age 14 Math Score: Between-sibship | -0.179 | 0.031 | 0.836 | [0.787, 0.888] | <0.001 | *** | 396466 | Main Model |
| SSB | 14 | Math | Sibling | Age 14 Math Score: Within-sibship | -0.104 | 0.044 | 0.901 | [0.827, 0.982] | 0.0197 | * | 396466 | Main Model |
| SSB | 14 | Math | Sibling | Sex | 0.784 | 0.053 | 2.19 | [1.974, 2.43] | <0.001 | *** | 396466 | Main Model |
| SSB | 14 | Math | Sibling | Birth Year | -0.13 | 0.008 | 0.878 | [0.864, 0.892] | <0.001 | *** | 396466 | Main Model |
| SSB | 14 | Math | Sibling | Birth Order | 0.002 | 0.027 | 1.002 | [0.95, 1.056] | 0.9485 | | 396466 | Main Model |
| SSB | 14 | Reading | Population | Age 14 Reading Score | 0.01 | 0.018 | 1.01 | [0.975, 1.046] | 0.5896 | | 853604 | Main Model |
| SSB | 14 | Reading | Population | Sex | 0.835 | 0.038 | 2.305 | [2.139, 2.483] | <0.001 | *** | 853604 | Main Model |
| SSB | 14 | Reading | Population | Birth Year | -0.125 | 0.005 | 0.882 | [0.874, 0.891] | <0.001 | *** | 853604 | Main Model |
| SSB | 14 | Reading | Population | Birth Order | 0.111 | 0.016 | 1.117 | [1.083, 1.153] | <0.001 | *** | 853604 | Main Model |
| SSB | 14 | Reading | Sibling | Age 14 Reading Score: Between-sibship | -0.009 | 0.032 | 0.991 | [0.931, 1.055] | 0.7880 | | 395032 | Main Model |
| SSB | 14 | Reading | Sibling | Age 14 Reading Score: Within-sibship | 0.054 | 0.046 | 1.055 | [0.964, 1.155] | 0.2370 | | 395032 | Main Model |
| SSB | 14 | Reading | Sibling | Sex | 0.806 | 0.053 | 2.239 | [2.018, 2.484] | <0.001 | *** | 395032 | Main Model |
| SSB | 14 | Reading | Sibling | Birth Year | -0.129 | 0.008 | 0.879 | [0.865, 0.893] | <0.001 | *** | 395032 | Main Model |
| SSB | 14 | Reading | Sibling | Birth Order | 0.017 | 0.027 | 1.017 | [0.965, 1.072] | 0.5275 | | 395032 | Main Model |

**Supplementary Table 10. Full regression results for analysis 2 examining the association between academic performance and bipolar disorder diagnosis, adjusted for prior scores.**

*Estimates from age 13 are corrected for age 10, and estimates for age 14 are corrected for ages 10 and 13. Estimates for age 10 are the same as the results presented in Supplementary Table 9 since they are not adjusted for any previous scores. SSB = Statistics Norway; SE = Standard Error; CI = Confidence Interval.*
**p < .05. **p < .01. ***p < .001.*

| Cohort | Age | Test | Model | Variable | Estimate | SE | HR | 95% CI | p | Significance | N | Analysis |
|---|---|---|---|---|---|---|---|---|---|---|---|---|
| SSB | 10 | Total Score | Population | Age 10 Total Score | 0.039 | 0.019 | 1.04 | [1.002, 1.079] | 0.0449 | * | 768505 | Adjusted Model |
| SSB | 10 | Total Score | Population | Sex | 0.863 | 0.041 | 2.37 | [2.187, 2.569] | <0.001 | *** | 768505 | Adjusted Model |
| SSB | 10 | Total Score | Population | Birth Year | -0.14 | 0.006 | 0.869 | [0.859, 0.88] | <0.001 | *** | 768505 | Adjusted Model |
| SSB | 10 | Total Score | Population | Birth Order | 0.121 | 0.017 | 1.129 | [1.092, 1.167] | <0.001 | *** | 768505 | Adjusted Model |
| SSB | 10 | Total Score | Sibling | Age 10 Total Score: Between-sibship | -0.001 | 0.035 | 0.999 | [0.933, 1.07] | 0.9788 | | 343898 | Adjusted Model |
| SSB | 10 | Total Score | Sibling | Age 10 Total Score: Within-sibship | 0.109 | 0.048 | 1.115 | [1.015, 1.225] | 0.0241 | * | 343898 | Adjusted Model |
| SSB | 10 | Total Score | Sibling | Sex | 0.824 | 0.06 | 2.28 | [2.027, 2.564] | <0.001 | *** | 343898 | Adjusted Model |
| SSB | 10 | Total Score | Sibling | Birth Year | -0.143 | 0.009 | 0.867 | [0.852, 0.882] | <0.001 | *** | 343898 | Adjusted Model |
| SSB | 10 | Total Score | Sibling | Birth Order | 0.004 | 0.032 | 1.004 | [0.943, 1.069] | 0.9115 | | 343898 | Adjusted Model |
| SSB | 10 | Math | Population | Age 10 Math Score | -0.022 | 0.019 | 0.978 | [0.942, 1.015] | 0.2396 | | 791751 | Adjusted Model |
| SSB | 10 | Math | Population | Sex | 0.864 | 0.041 | 2.373 | [2.189, 2.571] | <0.001 | *** | 791751 | Adjusted Model |
| SSB | 10 | Math | Population | Birth Year | -0.14 | 0.006 | 0.869 | [0.859, 0.88] | <0.001 | *** | 791751 | Adjusted Model |
| SSB | 10 | Math | Population | Birth Order | 0.118 | 0.017 | 1.125 | [1.088, 1.163] | <0.001 | *** | 791751 | Adjusted Model |
| SSB | 10 | Math | Sibling | Age 10 Math Score: Between-sibship | -0.089 | 0.035 | 0.915 | [0.854, 0.98] | 0.0106 | * | 359395 | Adjusted Model |
| SSB | 10 | Math | Sibling | Age 10 Math Score: Within-sibship | 0.051 | 0.046 | 1.052 | [0.962, 1.152] | 0.2655 | | 359395 | Adjusted Model |
| SSB | 10 | Math | Sibling | Sex | 0.812 | 0.058 | 2.252 | [2.01, 2.524] | <0.001 | *** | 359395 | Adjusted Model |
| SSB | 10 | Math | Sibling | Birth Year | -0.142 | 0.009 | 0.868 | [0.852, 0.883] | <0.001 | *** | 359395 | Adjusted Model |
| SSB | 10 | Math | Sibling | Birth Order | -0.002 | 0.031 | 0.998 | [0.939, 1.061] | 0.9407 | | 359395 | Adjusted Model |
| SSB | 10 | Reading | Population | Age 10 Reading Score | 0.126 | 0.02 | 1.134 | [1.091, 1.18] | <0.001 | *** | 779890 | Adjusted Model |
| SSB | 10 | Reading | Population | Sex | 0.848 | 0.041 | 2.335 | [2.155, 2.53] | <0.001 | *** | 779890 | Adjusted Model |

| | | | | | | | | | | | | |
|---|---|---|---|---|---|---|---|---|---|---|---|---|
| SSB | 10 | Reading | Population | Birth Year | -0.14 | 0.006 | 0.869 | [0.859, 0.88] | <0.001 | *** | 779890 | Adjusted Model |
| SSB | 10 | Reading | Population | Birth Order | 0.124 | 0.017 | 1.132 | [1.095, 1.17] | <0.001 | *** | 779890 | Adjusted Model |
| SSB | 10 | Reading | Sibling | Age 10 Reading Score: Between-sibship | 0.128 | 0.036 | 1.137 | [1.059, 1.22] | <0.001 | *** | 351087 | Adjusted Model |
| SSB | 10 | Reading | Sibling | Age 10 Reading Score: Within-sibship | 0.154 | 0.046 | 1.166 | [1.066, 1.277] | <0.001 | *** | 351087 | Adjusted Model |
| SSB | 10 | Reading | Sibling | Sex | 0.818 | 0.059 | 2.266 | [2.019, 2.544] | <0.001 | *** | 351087 | Adjusted Model |
| SSB | 10 | Reading | Sibling | Birth Year | -0.145 | 0.009 | 0.865 | [0.85, 0.88] | <0.001 | *** | 351087 | Adjusted Model |
| SSB | 10 | Reading | Sibling | Birth Order | 0.004 | 0.032 | 1.004 | [0.943, 1.069] | 0.8932 | | 351087 | Adjusted Model |
| SSB | 10 | English | Population | Age 10 English Score | 0.13 | 0.021 | 1.139 | [1.093, 1.187] | <0.001 | *** | 733986 | Adjusted Model |
| SSB | 10 | English | Population | Sex | 0.868 | 0.043 | 2.382 | [2.19, 2.592] | <0.001 | *** | 733986 | Adjusted Model |
| SSB | 10 | English | Population | Birth Year | -0.14 | 0.006 | 0.869 | [0.859, 0.88] | <0.001 | *** | 733986 | Adjusted Model |
| SSB | 10 | English | Population | Birth Order | 0.125 | 0.018 | 1.133 | [1.094, 1.174] | <0.001 | *** | 733986 | Adjusted Model |
| SSB | 10 | English | Sibling | Age 10 English Score: Between-sibship | 0.152 | 0.041 | 1.164 | [1.074, 1.262] | <0.001 | *** | 313825 | Adjusted Model |
| SSB | 10 | English | Sibling | Age 10 English Score: Within-sibship | 0.077 | 0.051 | 1.08 | [0.977, 1.194] | 0.1269 | | 313825 | Adjusted Model |
| SSB | 10 | English | Sibling | Sex | 0.806 | 0.065 | 2.239 | [1.971, 2.543] | <0.001 | *** | 313825 | Adjusted Model |
| SSB | 10 | English | Sibling | Birth Year | -0.147 | 0.009 | 0.863 | [0.848, 0.879] | <0.001 | *** | 313825 | Adjusted Model |
| SSB | 10 | English | Sibling | Birth Order | 0.036 | 0.033 | 1.037 | [0.972, 1.106] | 0.2754 | | 313825 | Adjusted Model |
| SSB | 13 | Total Score | Population | Age 13 Total Score | -0.168 | 0.037 | 0.845 | [0.786, 0.909] | <0.001 | *** | 728740 | Adjusted Model |
| SSB | 13 | Total Score | Population | Age 10 Total Score | 0.18 | 0.036 | 1.197 | [0.116, 0.285] | <0.001 | *** | 728740 | Adjusted Model |
| SSB | 13 | Total Score | Population | Sex | 0.907 | 0.044 | 2.477 | [0.272, 0.7] | <0.001 | *** | 728740 | Adjusted Model |
| SSB | 13 | Total Score | Population | Birth Year | -0.138 | 0.006 | 0.871 | [0.861, 0.881] | <0.001 | *** | 728740 | Adjusted Model |
| SSB | 13 | Total Score | Population | Birth Order | 0.122 | 0.018 | 1.13 | [0.091, 0.17] | <0.001 | *** | 728740 | Adjusted Model |
| SSB | 13 | Total Score | Sibling | Age 13 Total Score: Between-sibship | -0.247 | 0.061 | 0.781 | [0.693, 0.88] | <0.001 | *** | 320342 | Adjusted Model |
| SSB | 13 | Total Score | Sibling | Age 13 Total Score: Within-sibship | -0.173 | 0.07 | 0.841 | [0.733, 0.965] | 0.014 | * | 320342 | Adjusted Model |
| SSB | 13 | Total Score | Sibling | Age 10 Total Score | 0.226 | 0.055 | 1.254 | [1.125, 1.396] | <0.001 | *** | 320342 | Adjusted Model |
| SSB | 13 | Total Score | Sibling | Sex | 0.847 | 0.063 | 2.333 | [2.062, 2.639] | <0.001 | *** | 320342 | Adjusted Model |

| | | | | | | | | | | | | |
|---|---|---|---|---|---|---|---|---|---|---|---|---|
| SSB | 13 | Total Score | Sibling | Birth Year | -0.144 | 0.009 | 0.866 | [0.851, 0.881] | <0.001 | *** | 320342 | Adjusted Model |
| SSB | 13 | Total Score | Sibling | Birth Order | 0.016 | 0.034 | 1.016 | [0.951, 1.086] | 0.6365 | | 320342 | Adjusted Model |
| SSB | 13 | Math | Population | Age13 Math Score | -0.176 | 0.032 | 0.839 | [0.788, 0.893] | <0.001 | *** | 762117 | Adjusted Model |
| SSB | 13 | Math | Population | Age 10 Math Score | 0.114 | 0.031 | 1.121 | [1.055, 1.191] | <0.001 | *** | 762117 | Adjusted Model |
| SSB | 13 | Math | Population | Sex | 0.887 | 0.042 | 2.428 | [2.236, 2.636] | <0.001 | *** | 762117 | Adjusted Model |
| SSB | 13 | Math | Population | Birth Year | -0.14 | 0.006 | 0.869 | [0.859, 0.88] | <0.001 | *** | 762117 | Adjusted Model |
| SSB | 13 | Math | Population | Birth Order | 0.122 | 0.017 | 1.13 | [1.093, 1.168] | <0.001 | *** | 762117 | Adjusted Model |
| SSB | 13 | Math | Sibling | Age 13 Math Score: Between-sibship | -0.25 | 0.053 | 0.779 | [0.702, 0.864] | <0.001 | *** | 342070 | Adjusted Model |
| SSB | 13 | Math | Sibling | Age 13 Math Score: Within-sibship | -0.13 | 0.06 | 0.878 | [0.781, 0.988] | 0.0307 | * | 342070 | Adjusted Model |
| SSB | 13 | Math | Sibling | Age 10 Math Score | 0.132 | 0.046 | 1.141 | [1.043, 1.249] | 0.0046 | ** | 342070 | Adjusted Model |
| SSB | 13 | Math | Sibling | Sex | 0.823 | 0.061 | 2.277 | [2.021, 2.567] | <0.001 | *** | 342070 | Adjusted Model |
| SSB | 13 | Math | Sibling | Birth Year | -0.143 | 0.009 | 0.867 | [0.852, 0.882] | <0.001 | *** | 342070 | Adjusted Model |
| SSB | 13 | Math | Sibling | Birth Order | 0.007 | 0.032 | 1.007 | [0.946, 1.072] | 0.8323 | | 342070 | Adjusted Model |
| SSB | 13 | Reading | Population | Age 13 Reading Score | -0.057 | 0.031 | 0.945 | [0.889, 1.004] | 0.0619 | | 749640 | Adjusted Model |
| SSB | 13 | Reading | Population | Age 10 Reading Score | 0.177 | 0.03 | 1.194 | [1.125, 1.266] | <0.001 | *** | 749640 | Adjusted Model |
| SSB | 13 | Reading | Population | Sex | 0.865 | 0.043 | 2.375 | [2.183, 2.584] | <0.001 | *** | 749640 | Adjusted Model |
| SSB | 13 | Reading | Population | Birth Year | -0.138 | 0.006 | 0.871 | [0.861, 0.881] | <0.001 | *** | 749640 | Adjusted Model |
| SSB | 13 | Reading | Population | Birth Order | 0.122 | 0.018 | 1.13 | [1.091, 1.17] | <0.001 | *** | 749640 | Adjusted Model |
| SSB | 13 | Reading | Sibling | Age 13 Reading Score: Between-sibship | -0.123 | 0.052 | 0.884 | [0.799, 0.979] | 0.0195 | * | 333201 | Adjusted Model |
| SSB | 13 | Reading | Sibling | Age 13 Reading Score: Within-sibship | -0.076 | 0.06 | 0.927 | [0.824, 1.042] | 0.2031 | | 333201 | Adjusted Model |
| SSB | 13 | Reading | Sibling | Age 10 Reading Score | 0.224 | 0.046 | 1.251 | [1.143, 1.369] | <0.001 | *** | 333201 | Adjusted Model |
| SSB | 13 | Reading | Sibling | Sex | 0.821 | 0.062 | 2.273 | [2.013, 2.566] | <0.001 | *** | 333201 | Adjusted Model |
| SSB | 13 | Reading | Sibling | Birth Year | -0.145 | 0.009 | 0.865 | [0.85, 0.88] | <0.001 | *** | 333201 | Adjusted Model |
| SSB | 13 | Reading | Sibling | Birth Order | 0.015 | 0.033 | 1.015 | [0.952, 1.083] | 0.6438 | | 333201 | Adjusted Model |
| SSB | 13 | English | Population | Age 13 English Score | 0.159 | 0.032 | 1.172 | [1.101, 1.248] | <0.001 | *** | 702751 | Adjusted Model |

| | | | | | | | | | | | | |
|---|---|---|---|---|---|---|---|---|---|---|---|---|
| SSB | 13 | English | Population | Age 10 English Score | 0.028 | 0.031 | 1.028 | [0.968, 1.093] | 0.363 | | 702751 | Adjusted Model |
| SSB | 13 | English | Population | Sex | 0.897 | 0.044 | 2.452 | [2.25, 2.673] | <0.001 | *** | 702751 | Adjusted Model |
| SSB | 13 | English | Population | Birth Year | -0.139 | 0.006 | 0.87 | [0.86, 0.881] | <0.001 | *** | 702751 | Adjusted Model |
| SSB | 13 | English | Population | Birth Order | 0.124 | 0.019 | 1.132 | [1.091, 1.175] | <0.001 | *** | 702751 | Adjusted Model |
| SSB | 13 | English | Sibling | Age 13 English Score: Between-sibship | 0.21 | 0.055 | 1.234 | [1.108, 1.374] | <0.001 | *** | 295865 | Adjusted Model |
| SSB | 13 | English | Sibling | Age 13 English Score: Within-sibship | 0.113 | 0.065 | 1.12 | [0.986, 1.272] | 0.0822 | | 295865 | Adjusted Model |
| SSB | 13 | English | Sibling | Age 10 English Score | 0.009 | 0.047 | 1.009 | [0.92, 1.106] | 0.8449 | | 295865 | Adjusted Model |
| SSB | 13 | English | Sibling | Sex | 0.829 | 0.068 | 2.291 | [2.005, 2.618] | <0.001 | *** | 295865 | Adjusted Model |
| SSB | 13 | English | Sibling | Birth Year | -0.151 | 0.01 | 0.86 | [0.843, 0.877] | <0.001 | *** | 295865 | Adjusted Model |
| SSB | 13 | English | Sibling | Birth Order | 0.049 | 0.034 | 1.05 | [0.983, 1.123] | 0.157 | | 295865 | Adjusted Model |
| SSB | 14 | Total Score | Population | Age 14 Total Score | -0.409 | 0.049 | 0.664 | [0.603, 0.731] | <0.001 | *** | 693836 | Adjusted Model |
| SSB | 14 | Total Score | Population | Age 13 Total Score | 0.248 | 0.039 | 1.281 | [1.187, 1.383] | <0.001 | *** | 693836 | Adjusted Model |
| SSB | 14 | Total Score | Population | Age 10 Total Score | 0.148 | 0.054 | 1.16 | [1.043, 1.289] | 0.0057 | ** | 693836 | Adjusted Model |
| SSB | 14 | Total Score | Population | Sex | 0.888 | 0.045 | 2.43 | [2.225, 2.654] | <0.001 | *** | 693836 | Adjusted Model |
| SSB | 14 | Total Score | Population | Birth Year | -0.142 | 0.007 | 0.868 | [0.856, 0.88] | <0.001 | *** | 693836 | Adjusted Model |
| SSB | 14 | Total Score | Population | Birth Order | 0.122 | 0.019 | 1.13 | [1.088, 1.173] | <0.001 | *** | 693836 | Adjusted Model |
| SSB | 14 | Total Score | Sibling | Age 14 Total Score: Between-sibship | -0.439 | 0.083 | 0.645 | [0.548, 0.759] | <0.001 | *** | 299039 | Adjusted Model |
| SSB | 14 | Total Score | Sibling | Age 14 Total Score: Within-sibship | -0.33 | 0.086 | 0.719 | [0.607, 0.851] | <0.001 | *** | 299039 | Adjusted Model |
| SSB | 14 | Total Score | Sibling | Age 13 Total Score | 0.241 | 0.06 | 1.273 | [1.131, 1.431] | <0.001 | *** | 299039 | Adjusted Model |
| SSB | 14 | Total Score | Sibling | Age 10 Total Score | 0.159 | 0.083 | 1.172 | [0.996, 1.379] | 0.056 | | 299039 | Adjusted Model |
| SSB | 14 | Total Score | Sibling | Sex | 0.821 | 0.067 | 2.273 | [1.993, 2.592] | <0.001 | *** | 299039 | Adjusted Model |
| SSB | 14 | Total Score | Sibling | Birth Year | -0.149 | 0.01 | 0.862 | [0.845, 0.879] | <0.001 | *** | 299039 | Adjusted Model |
| SSB | 14 | Total Score | Sibling | Birth Order | 0.032 | 0.035 | 1.033 | [0.964, 1.106] | 0.3594 | | 299039 | Adjusted Model |
| SSB | 14 | Math | Population | Age 14 Math Score | -0.31 | 0.043 | 0.733 | [0.674, 0.798] | <0.001 | *** | 734383 | Adjusted Model |
| SSB | 14 | Math | Population | Age 13 Math Score | 0.171 | 0.033 | 1.186 | [1.112, 1.266] | <0.001 | *** | 734383 | Adjusted Model |
| SSB | 14 | Math | Population | Age 10 Math Score | 0.06 | 0.045 | 1.062 | [1.972, 1.16] | 0.1883 | | 734383 | Adjusted Model |

| | | | | | | | | | | | | |
|---|---|---|---|---|---|---|---|---|---|---|---|---|
| SSB | 14 | Math | Population | Sex | 0.855 | 0.043 | 2.351 | [2.161, 2.558] | <0.001 | *** | 734383 | Adjusted Model |
| SSB | 14 | Math | Population | Birth Year | -0.143 | 0.007 | 0.867 | [0.855, 0.879] | <0.001 | *** | 734383 | Adjusted Model |
| SSB | 14 | Math | Population | Birth Order | 0.121 | 0.018 | 1.129 | [1.09, 1.169] | <0.001 | *** | 734383 | Adjusted Model |
| SSB | 14 | Math | Sibling | Age 14 Math Score: Between-sibship | -0.355 | 0.072 | 0.701 | [0.609, 0.807] | <0.001 | *** | 324898 | Adjusted Model |
| SSB | 14 | Math | Sibling | Age 14 Math Score: Within-sibship | -0.251 | 0.076 | 0.778 | [0.67, 0.903] | <0.001 | *** | 324898 | Adjusted Model |
| SSB | 14 | Math | Sibling | Age 13 Math Score | 0.171 | 0.05 | 1.186 | [1.076, 1.309] | <0.001 | *** | 324898 | Adjusted Model |
| SSB | 14 | Math | Sibling | Age 10 Math Score | 0.067 | 0.069 | 1.069 | [0.934, 1.224] | 0.3309 | | 324898 | Adjusted Model |
| SSB | 14 | Math | Sibling | Sex | 0.794 | 0.064 | 2.212 | [1.951, 2.508] | <0.001 | *** | 324898 | Adjusted Model |
| SSB | 14 | Math | Sibling | Birth Year | -0.146 | 0.01 | 0.864 | [0.847, 0.881] | <0.001 | *** | 324898 | Adjusted Model |
| SSB | 14 | Math | Sibling | Birth Order | 0.019 | 0.033 | 1.019 | [0.955, 1.087] | 0.5738 | | 324898 | Adjusted Model |
| SSB | 14 | Reading | Population | Age 14 Reading Score | -0.23 | 0.037 | 0.795 | [0.739, 0.854] | <0.001 | *** | 724413 | Adjusted Model |
| SSB | 14 | Reading | Population | Age 13 Reading Score | 0.226 | 0.032 | 1.254 | [1.177, 1.335] | <0.001 | *** | 724413 | Adjusted Model |
| SSB | 14 | Reading | Population | Age 10 Reading Score | 0.094 | 0.039 | 1.099 | [1.018, 1.186] | 0.0166 | * | 724413 | Adjusted Model |
| SSB | 14 | Reading | Population | Sex | 0.874 | 0.044 | 2.396 | [2.198, 2.612] | <0.001 | *** | 724413 | Adjusted Model |
| SSB | 14 | Reading | Population | Birth Year | -0.141 | 0.006 | 0.868 | [0.858, 0.879] | <0.001 | *** | 724413 | Adjusted Model |
| SSB | 14 | Reading | Population | Birth Order | 0.121 | 0.018 | 1.129 | [1.09, 1.169] | <0.001 | *** | 724413 | Adjusted Model |
| SSB | 14 | Reading | Sibling | Age 14 Reading Score: Between-sibship | -0.316 | 0.064 | 0.729 | [0.643, 0.826] | <0.001 | *** | 317546 | Adjusted Model |
| SSB | 14 | Reading | Sibling | Age 14 Reading Score: Within-sibship | -0.154 | 0.068 | 0.857 | [0.75, 0.979] | 0.0228 | * | 317546 | Adjusted Model |
| SSB | 14 | Reading | Sibling | Age 13 Reading Score | 0.266 | 0.05 | 1.305 | [0.183, 0.439] | <0.001 | *** | 317546 | Adjusted Model |
| SSB | 14 | Reading | Sibling | Age 10 Reading Score | 0.098 | 0.06 | 1.103 | [0.981, 0.241] | 0.1029 | | 317546 | Adjusted Model |
| SSB | 14 | Reading | Sibling | Sex | 0.815 | 0.065 | 2.259 | [0.989, 0.566] | <0.001 | *** | 317546 | Adjusted Model |
| SSB | 14 | Reading | Sibling | Birth Year | -0.146 | 0.01 | 0.864 | [0.847, 0.881] | <0.001 | *** | 317546 | Adjusted Model |
| SSB | 14 | Reading | Sibling | Birth Order | 0.024 | 0.034 | 1.024 | [0.958, 0.095] | 0.4701 | | 317546 | Adjusted Model |

**Supplementary Table 11. Statistical tests for non-linearity in the association between academic performance and bipolar disorder diagnosis.**

*p values are from likelihood-ratio tests comparing Cox proportional hazards models with school test performance modelled as a linear continuous predictor with models in which school test performance was modelled using a natural cubic spline with four degrees of freedom. A significant p value indicates that the non-linear association is significantly different from linear models. SSB = Statistics Norway; CBS = Statistics Netherlands.*

*$^*p < .05$. $^{**}p < .01$. $^{***}p < .001$.*

| Cohort | Age | Test | N | $\chi^2$ | df | p | Significance |
|---|---|---|---|---|---|---|---|
| SSB | 10 | Total Score | 768505 | 1.247 | 3 | 0.742 | |
| SSB | 10 | Math | 791751 | 8.746 | 3 | 0.033 | * |
| SSB | 10 | Reading | 779890 | 3.327 | 3 | 0.344 | |
| SSB | 10 | English | 733986 | 12.271 | 3 | 0.006 | * |
| CBS | 12 | Total Score | 1144287 | 1.857 | 3 | 0.602 | |
| CBS | 12 | Math | 1144196 | 0.311 | 3 | 0.958 | |
| CBS | 12 | Reading | 1142246 | 7.782 | 3 | 0.051 | |
| CBS | 12 | Language | 1144196 | 2.799 | 3 | 0.424 | |
| SSB | 13 | Total Score | 949575 | 6.6 | 3 | 0.086 | |
| SSB | 13 | Math | 976455 | 1.926 | 3 | 0.588 | |
| SSB | 13 | Reading | 969899 | 0.275 | 3 | 0.964 | |
| SSB | 13 | English | 971859 | 4.697 | 3 | 0.195 | |
| SSB | 14 | Total Score | 833916 | 2.003 | 3 | 0.572 | |
| SSB | 14 | Math | 855298 | 1.695 | 3 | 0.638 | |
| SSB | 14 | Reading | 853604 | 2.847 | 3 | 0.416 | |

**Supplementary Table 12. Classes of Prescriptions used in the Prediction Model, their English translation and corresponding regression coefficients.**

*The prescription classes are based on the ATC (Anatomical Therapeutic Chemical) system. SE = Standard Error; OR = Odds Ratio; CI = Confidence Interval.*

*$*p < .05$. $**p < .01$. $***p < .001$.*

| ATC Codes | Class of Drug | | Estimate | SE | OR | 95% CI | p | Significance | N |
|---|---|---|---|---|---|---|---|---|---|
| | Dutch | English Translation | | | | | | | |
| | Intercept | | 0.246 | 0.017 | 1.279 | [1.237, 1.322] | <0.001 | *** | 32427 |
| N01A | Algemene anaesthetica | General anaesthetics | -0.136 | 0.402 | 0.873 | [0.40, 1.92] | 0.7358 | | 32427 |
| N01B | Anaesthetica, lokale | Anaesthetics, local | -0.325 | 0.109 | 0.723 | [0.583, 0.896] | 0.003 | ** | 32427 |
| N02A | Opioiden | Opioids | -0.443 | 0.046 | 0.642 | [0.586, 0.703] | <0.001 | *** | 32427 |
| N02B | Overige analgetica en antipyretica | Other analgesics and antipyretics | -0.401 | 0.082 | 0.670 | [0.57, 0.787] | <0.001 | *** | 32427 |
| N02C | Migrainemiddelen | Migraine drugs | -0.306 | 0.092 | 0.736 | [0.614, 0.883] | <0.001 | *** | 32427 |
| N03A | Anti-epileptica | Antiepileptics | 1.332 | 0.048 | 3.789 | [3.449, 4.165] | <0.001 | *** | 32427 |
| N04A | Parasympathicolytica | Parasympatholytics | 0.047 | 0.167 | 1.048 | [0.755, 1.454] | 0.7793 | | 32427 |
| N04B | Dopaminergica | Dopaminergic | -0.414 | 0.131 | 0.661 | [0.511, 0.855] | 0.0016 | ** | 32427 |
| N05A | Antipsychotica | Antipsychotics | 1.739 | 0.033 | 5.692 | [5.333, 6.075] | <0.001 | *** | 32427 |
| N05B | Anxiolytica | Anxiolytics | 0.116 | 0.043 | 1.123 | [1.031, 1.223] | 0.0078 | ** | 32427 |
| N05C | Hypnotica en sedativa | Hypnotics and sedatives | 0.278 | 0.051 | 1.320 | [1.196, 1.458] | <0.001 | *** | 32427 |
| N06A | Antidepressiva | Antidepressants | -0.617 | 0.033 | 0.540 | [0.505, 0.576] | <0.001 | *** | 32427 |
| N06B | Psychostimul., midd adhd/nootropica | Psychostimulants, ADHD drugs/nootropics | -0.516 | 0.071 | 0.597 | [0.519, 0.686] | <0.001 | *** | 32427 |
| N06C | Psycholeptica met psychoanaleptica | Psycholeptics with psychoanaleptics | - | - | - | - | - | | - |
| N06D | Dementiemiddelen | Dementia drugs | -0.245 | 0.376 | 0.783 | [0.374, 1.636] | 0.5142 | | 32427 |
| N07A | Parasympathicomimetica | Parasympathicomimetics | -0.914 | 0.528 | 0.401 | [0.142, 1.128] | 0.0832 | | 32427 |
| N07B | Middelen bij verslavingen | Drugs for addictions | -0.495 | 0.085 | 0.610 | [0.516, 0.72] | <0.001 | *** | 32427 |
| N07C | Vertigomiddelen | Vertigo drugs | -0.643 | 0.242 | 0.526 | [0.327, 0.846] | 0.008 | ** | 32427 |
| N07X | Ov. midd. werkzaam op zenuwstelsel | Other drugs active on the nervous system | -0.721 | 0.407 | 0.486 | [0.219, 1.079] | 0.0762 | | 32427 |

**Supplementary Table 13. Predictive performance of the bipolar disorder prediction model at different probability thresholds in the validation dataset.**

*The predicted probability threshold represents the minimum predicted probability used to classify an individual as a predicted case; individuals with probabilities below the threshold were classified as predicted controls. TP (true positive) = individuals correctly classified as cases; FP (false positive) = individuals classified as cases who were not observed cases; TN (true negative) = individuals correctly classified as controls; FN (false negative) = individuals classified as controls who were observed cases. Sensitivity = proportion of observed cases correctly identified by the prediction model [TP/(TP+FN)]; specificity = proportion of observed controls correctly identified [TN/(TN+FP)]; PPV (positive predictive value) = proportion of predicted cases who were observed cases [TP/(TP+FP)]; NPV (negative predictive value) = proportion of predicted controls who were observed controls [TN/(TN+FN)].*

| Probability Threshold | True Positives (TP) | False Positives (FP) | True Negatives (TN) | False Negatives (FN) | Sensitivity | Specificity | PPV | NPV |
|---|---|---|---|---|---|---|---|---|
| | | | | | TP/(TP + FN) | TN/(TN + FP) | TP/(TP + FP) | TN/(TN + FN) |
| 0.6 | 3605 | 520 | 1819 | 2163 | 0.625 | 0.778 | 0.874 | 0.457 |
| 0.7 | 3510 | 472 | 1867 | 2258 | 0.608 | 0.798 | 0.881 | 0.453 |
| 0.8 | 2889 | 314 | 2025 | 2879 | 0.501 | 0.865 | 0.902 | 0.413 |
| 0.9 | 1201 | 70 | 2269 | 4567 | 0.208 | 0.97 | 0.945 | 0.332 |

**Supplementary References**

1 KING: Relationship Inference Software. https://www.kingrelatedness.com/ (accessed July 20, 2026).

2 Manichaikul A, Mychaleckyj JC, Rich SS, Daly K, Sale M, Chen W-M. Robust relationship inference in genome-wide association studies. *Bioinformatics* 2010; **26**: 2867–73.

3 Cheesman R, Coleman J, Rayner C, *et al.* Familial Influences on Neuroticism and Education in the UK Biobank. *Behav Genet* 2020; **50**: 84–93.

4 Bycroft C, Freeman C, Petkova D, *et al.* The UK Biobank resource with deep phenotyping and genomic data. *Nature* 2018; **562**: 203–9.

5 Auton A, Abecasis GR, Altshuler DM, *et al.* A global reference for human genetic variation. *Nature* 2015; **526**: 68–74.

6 Abdellaoui A, Hugh-Jones D, Yengo L, *et al.* Genetic correlates of social stratification in Great Britain. *Nat Hum Behav* 2019; **3**: 1332–42.

7 Amin V, Fletcher JM, Sun Z, Lu Q. Higher educational attainment is associated with longer telomeres in midlife: Evidence from sibling comparisons in the UK Biobank. *SSM - Population Health* 2022; **17**: 101018.

8 Carter AR, Harrison S, Gill D, *et al.* Educational attainment as a modifier for the effect of polygenic scores for cardiovascular risk factors: cross-sectional and prospective analysis of UK Biobank. *International Journal of Epidemiology* 2022; **51**: 885–97.

9 Saunes IS, Karanikolos M, Sagan A. Norway: Health System Review. *Health Syst Transit* 2020; **22**: 1–163.

10 Okbay A, Wu Y, Wang N, *et al.* Polygenic prediction of educational attainment within and between families from genome-wide association analyses in 3 million individuals. *Nat Genet* 2022; **54**: 437–49.

11 Mullins N, Forstner AJ, O'Connell KS, *et al.* Genome-wide association study of more than 40,000 bipolar disorder cases provides new insights into the underlying biology. *Nat Genet* 2021; **53**: 817–29.

12 Kendler KS, Ohlsson H, Mezuk B, Sundquist K, Sundquist J. A Swedish National Prospective and Co-relative Study of School Achievement at Age 16, and Risk for Schizophrenia, Other Nonaffective Psychosis, and Bipolar Illness. *Schizophrenia Bulletin* 2016; **42**: 77–86.

13 MacCabe JH, Lambe MP, Cnattingius S, *et al.* Excellent school performance at age 16 and risk of adult bipolar disorder: national cohort study. *Br J Psychiatry* 2010; **196**: 109–15.

14 predict function - RDocumentation. https://www.rdocumentation.org/packages/stats/versions/3.6.2/topics/predict (accessed Aug 19, 2026).
